# Stage–Specific Roles of Plasma GFAP in Linking Amyloid Cascade in Alzheimer Disease

**DOI:** 10.64898/2026.11.03.26366374

**Authors:** Xuan Niu, Qiuchang Sun, Julie K. Wisch, Meng Jiang, Aime Luna, David A. Hoagey, Suzanne E. Schindler, Carlos Cruchaga, Brian A. Gordon, Yong Wang, Tammie L.S. Benzinger, Alzheimer’s Disease Neuroimaging Initiative (ADNI), Qing Wang

## Abstract

**INTRODUCTION:** Astrocytic reactivity, measured by plasma glial fibrillary acidic protein (GFAP), is robustly elevated across the Alzheimer disease (AD) continuum, yet its relationship with the amyloid cascade remains poorly understood.

**METHODS:** We integrated plasma GFAP with multimodal AD biomarkers including cerebrospinal fluid (CSF) or plasma Aβ (Aβ42/40), Aβ and tau positron emission tomography (PET), structural MRI, and clinical severity, across a discovery cohort (Knight Alzheimer Disease Research Center, n=278) and an independent replication cohort (Alzheimer’s Disease Neuroimaging Initiative, n=236).

**RESULTS:** In both cohorts, plasma GFAP mediated the association of soluble Aβ42/40 (CSF in the Knight ADRC; plasma in ADNI) with Aβ PET, as well as the association of Aβ PET with tau PET in the overall samples, which remained significant after false discovery rate (FDR) correction. Among tau-negative individuals, GFAP mediated the association between CSF Aβ42/40 and Aβ PET in Knight ADRC, but this effect did not survive FDR correction with plasma Aβ42/40 in ADNI. In Aβ-positive individuals, GFAP mediated the association between Aβ PET and tau PET burden in both cohorts, but survived correction only in ADNI. Interaction findings were partially replicated across cohorts. Exploratory plasma proteomics further revealed a GFAP-associated protein network enriched for extracellular matrix (ECM)-immune remodeling network.

**DISCUSSION:** These findings reveal stage-specific associations of plasma GFAP with the amyloid cascade and implicate an astrocyte-centered ECM–inflammatory axis, which may inform future diagnostic panels or targeted therapies.

**Clinical trial registration:** Alzheimer’s Disease Neuroimaging Initiative 3, ClinicalTrials.gov, NCT02854033.

## 1 BACKGROUND

Alzheimer disease (AD) is biologically characterized by the accumulation of amyloid-β (Aβ) and tau aggregates in the brain. The classical Aβ cascade hypothesis posits that Aβ accumulation is the primary upstream event that triggers tau deposition and ultimately leads to neurodegeneration and cognitive impairment. Accumulating evidence suggests that the Aβ cascade alone does not fully explain the clinical and biological progression of AD^1^, but rather involves complex interactions with astrocytic reactivity, positioning neuroinflammation as a third prominent feature that drives disease progression^2^. However, the specific role of astrocytic reactivity within the Aβ cascade across the AD continuum remains poorly defined.

Although plasma glial fibrillary acidic protein (GFAP) is recognized as a marker of astrogliosis associated with amyloid pathology^3^, emerging evidence further suggests that plasma GFAP mediates the relationship between Aβ and tau accumulation^4,5^. Notably, changes in soluble Aβ are detectable in cerebrospinal fluid (CSF) and plasma (for example, CSF or plasma Aβ42/40) years before fibrillar Aβ deposition becomes apparent on positron emission tomography (PET)^6,7^. Plasma GFAP has been found to rise early in the course of AD, correlating with continuous amyloid-PET signal even in cognitively unimpaired individuals with normal Aβ PET^3^. A similar temporal sequence has been observed in adults with Down syndrome, a genetically determined model of AD driven by lifelong amyloid precursor protein overexpression: plasma GFAP levels increase in parallel with CSF Aβ declines approximately a decade prior to increases in amyloid-PET uptake, and continue to rise across the AD continuum^8^. Together, these findings raise the possibility that astrocytic reactivity may influence not only the link from Aβ to tau pathology, but also the earlier association between soluble Aβ dysregulation and insoluble amyloid deposition. Moreover, converging evidence indicates that Aβ facilitates downstream tau phosphorylation and propagation, thereby accelerating disease progression ^9,10^. This tau pathology is, in turn, more closely associated with neuronal injury and cognitive decline than Aβ itself^11^. Whether the influence of astrocytic reactivity on the AD pathological cascade differs according to participants’ amyloid/tau (A/T) biomarker status remains unknown. To address this question and to probe the underlying biological mechanisms, we additionally leveraged plasma proteomics, which offers a powerful approach to identify pathways underlying the association between astrocytic reactivity and AD pathology and to highlight candidate therapeutic targets.

In this study, we focused on plasma GFAP as a marker of astrocytic reactivity and integrated it with multimodal AD biomarkers and large–scale plasma proteomics in a discovery cohort (Knight Alzheimer Disease Research Center [Knight ADRC]), with replication of the key findings in an independent cohort (Alzheimer’s Disease Neuroimaging Initiative [ADNI]). We investigated the associations of plasma GFAP with different components of the AD cascade in the overall sample, and within subgroups of individuals without detectable tau PET pathology or with established Aβ pathology. Further, we conducted pathway and network analyses of the plasma GFAP–associated proteomic module that overlapped with core Aβ and tau pathologies to identify key biological pathways and hub proteins that may link astrocytic activation to these pathological AD biomarkers.

## 2 MATERIALS AND METHODS

### 2.1 Study Participants

The study included participants from two cohorts: The Knight ADRC (data freeze 25, released November 30th, 2024), and the ADNI phase 3 (http://adni.loni.usc.edu/). The ADNI was launched in 2003 as a public-private partnership, led by Principal Investigator Michael W. Weiner, MD. For up-to-date information, see adni.loni.usc.edu. Participants from both cohorts who had available plasma GFAP, soluble Aβ measures (CSF Aβ42/40 in the Knight ADRC and plasma Aβ42/40 in ADNI3), Aβ PET, tau PET, and structural magnetic resonance imaging (MRI) within 1 year of clinical assessment were included. Participants completed clinic visits including assessment of clinical dementia rating (CDR^®^) and CDR Sum of Boxes (CDR-SB)^12^. Cognitive status was defined according to the global CDR score, with CDR = 0 classified as cognitively normal (CN) and CDR > 0 as cognitively impaired (CI). We assessed 514 individuals: 278 from the Knight ADRC cohort (229 CN, 49 CI), and 236 from the ADNI cohort (163 CN, 73 CI).

Among Knight ADRC participants, 252 individuals had available proteomics data. The present study complies with all relevant ethical regulations, in accordance with the Declaration of Helsinki. All protocols were approved by the Washington University in St. Louis institutional review board (Human Research Protection Office) and by the local institutional review boards at each participating ADNI site. Written informed consent was obtained from all participants or legally authorized representatives.

### 2.2 Fluid biomarkers

Plasma and CSF samples were collected as previously described in the Knight ADRC and ADNI cohorts^13^. In the Knight ADRC cohort, CSF Aβ40 and Aβ42 concentrations were quantified by chemiluminescent enzyme immunoassay using a fully automated platform (LUMIPULSE G1200, Fujirebio, Malvern, PA, USA). In the ADNI cohort, plasma Aβ40 and Aβ42 concentrations were measured using a validated liquid chromatography–tandem mass spectrometry (LC-MS/MS) method developed at Washington University School of Medicine, as previously described^14^. Plasma levels of GFAP were measured in both cohorts using commercially available Simoa Discovery immunoassay (Quanterix). All assays were performed blinded to clinical and imaging data.

### 2.3 Proteomics data and quality control

Plasma proteomic profiling was performed using the SomaScan v4 assay (SomaLogic Inc.), a multiplexed single–stranded DNA aptamer platform enabling quantification of approximately 7,000 protein targets^15^. Unfasted plasma samples were collected at the time of clinical visit, immediately centrifuged and stored at −80 °C^16^. No preprocessing was needed for plasma samples prior to shipping. SomaLogic conducted the initial measurements and data normalization to control for inter-plate and intra-plate variability using hybridization controls and median signal intensities, as well as biological variance using an external reference.

Subsequently, the GHTO team performed an in-depth quality control (QC) as described previously^17^. Briefly, scale factors were computed for each aptamer in each plate; aptamers with a scale factor difference greater than 0.5 between any pairwise plate comparison were considered unreliable and removed. Next, the coefficient of variation (CV) was calculated for each aptamer by plate, and aptamers with a CV difference greater than 0.15 in any pairwise comparison were flagged as low quality and excluded. For the remaining aptamers, outliers were identified based on inter-quartile range (IQR) thresholds, and aptamers with missing rates >65% and >85% at successive filtering steps were removed. Following QC process, protein levels were log₁₀-transformed, and proteins with a missing rate greater than 20% were excluded prior to downstream analysis.

### 2.4 Neuroimaging

#### 2.4.1 Structural MRI

T1-weighted scans were processed using FreeSurfer (Martinos Center for Biomedical Imaging, Charlestown, Massachusetts, USA), and the Desikan–Killiany atlas was applied. In the Knight ADRC, imaging was performed using either a 3 T Siemens Biograph mMR or TIM Trio (Erlangen, Germany) scanner. Scanning parameters can be found in **Supplementary Methods**. In the ADNI, structural MRI data were obtained on 3T scanners using unified scanning protocols (parameter details can be found on: https://adni.loni.usc.edu/wpcontent/uploads/2017/07/ADNI3-MRI-protocols.pdf). In both cohorts, total hippocampal volume (THV, mm^3^) was calculated across hemispheres from the structural MRI scans and corrected for intracranial volume (ICV) using the residual method described by Jack et al^18^.

#### 2.4.2 PET imaging

For Knight ADRC participants, Aβ PET imaging was performed using ¹¹C-Pittsburgh Compound B (PiB)^19^, ¹⁸F-Florbetapir (FBP)^20^, and ¹⁸F-Florbetaben (FBB)^21^. In the ADNI, Aβ PET imaging was performed using ¹⁸F-Florbetaben (FBB) and ¹⁸F-Florbetapir (FBP)^22,23^. We applied a previously published method to convert the Aβ PET standardized uptake value ratio (SUVR) to the Centiloid scale^24^. Amyloid SUVRs are calculated using the cerebellar cortex in the Knight ADRC and the whole cerebellum in the ADNI as the reference region, further scaled to a global Centiloid composite calculated from the mean lateral orbitofrontal, medial orbitofrontal, precuneus, rostral middle frontal, superior frontal, superior temporal, and middle temporal regions^24^. Aβ PET positivity was defined as Centiloid value equal or higher to 20, as previously described^25^.

^18^F-Flortaucipir (FTP) was used for in-vivo imaging of tau pathology in both Knight ADRC and ADNI cohorts. For tau PET, SUVRs were calculated using the cerebellar gray in the Knight ADRC and inferior cerebellar gray in the ADNI as the reference region. Cortical tau tangles were evaluated by using a predefined temporal meta–region of interest (temporal-MetaROI), including the entorhinal cortex, parahippocampal gyrus, amygdala, fusiform gyrus, inferior temporal and middle temporal brain regions. The tau positivity (T+) was defined as temporal-MetaROI ^18^FTP SUVR ≥ 1.3 as described previously, typically ranging between SUVR 1.2 and 1.4^26–28^.

### 2.5 Statistical analysis

Statistical analyses were carried out using R software (version 4.0.2, http://www.r-project.org/), unless stated otherwise. The A−T+ group was excluded from statistical analyses, reflecting non-AD pathologic change within the AT(N) framework. All analyses were adjusted for age, sex, and cognitive status, except those involving CDR-SB, which were adjusted for age and sex only.

Initially, partial Spearman’s correlation analyses were conducted in all participants to examine associations between plasma GFAP and different components of the AD cascade, including soluble Aβ measures (CSF Aβ42/40 in the Knight ADRC cohort and plasma Aβ42/40 in ADNI), Aβ PET (global Centiloid), tau PET (temporal-MetaROI SUVR), structural MRI (Hippocampal volume), and clinical severity (CDR-SB). To further investigate these associations across the AD continuum, participants were stratified according to biological staging by PET imaging (A−T−, A+T−, A+T+). Additional subgroup analyses were performed in T− individuals (A−T− and A+T−) to examine GFAP-related associations in the absence of detectable tau PET pathology, and in A+ individuals (A+T− and A+T+) to examine these associations in the presence of established amyloid pathology.

Mediation analyses were performed to test whether astrocytic reactivity (plasma GFAP) mediated the associations along the AD pathological cascade in all participants and stratified by A/T stages, including soluble Aβ → Aβ PET, Aβ PET → tau PET, tau PET → structural MRI/ clinical severity. Indirect effects were estimated using bias-corrected bootstrap confidence intervals (5,000 resamples), and mediation was considered significant if the 95% confidence interval did not include zero. In parallel, we assessed whether plasma GFAP modified the strength of associations between sequential pathological markers. Using linear regression models, interaction terms were tested at each step of the cascade (e.g., GFAP × soluble Aβ on AβPET; GFAP × Aβ PET on tau PET; GFAP × tau PET on structural MRI or clinical severity) in the overall sample, as well as in T− and A+ subgroups. Participants were dichotomized into two groups based on the median value of plasma GFAP: low-GFAP, comprising participants with GFAP levels in the lower half, and high-GFAP, comprising those with levels in the upper half.

When a significant interaction was detected (P < 0.05), we further performed stratified analyses to estimate the associations within each GFAP subgroup separately. Region–wise regression coefficients (β) from the interaction models and from the subsequent analyses stratified by low versus high plasma GFAP were estimated at the ROI level. To account for multiple comparisons, P values for the indirect effects (mediation analyses) and interaction terms were adjusted using the Benjamini–Hochberg false discovery rate (FDR) procedure within each cohort and analysis group (overall sample, T− subgroup, and A+ subgroup). The corrected P values are denoted as PFDR. Both unadjusted P values and PFDR values are reported. To further examine whether plasma GFAP modifies amyloid-related associations in the absence of tau pathology and at relatively lower levels of amyloid burden, we conducted sensitivity analyses restricted to tau-negative individuals with Aβ PET < 50 Centiloids. This cutoff was informed by a PET–autopsy study reporting that approximately 50 Centiloids best identified intermediate-to-high Alzheimer’s disease neuropathologic change (ADNC) ^29^. The analysis was repeated using nearby thresholds (< 46, < 48, < 52, < 54, and < 56 Centiloids). These sensitivity analyses were exploratory and were not corrected for multiple comparisons.

Finally, to further elucidate the role of astrocytic reactivity across the AD continuum, structural equation modeling (SEM) was conducted using the R package lavaan. Based on the proposed AD cascade and the subgroup findings, separate models were constructed for T− and A+ individuals: (1) in T− individuals: CSF Aβ42/40/plasma Aβ42/40 → plasma GFAP → Aβ PET → Tau PET → structural MRI/clinical severity; (2) A+ individuals: CSF Aβ42/40 or plasma Aβ42/40 → Aβ PET → plasma GFAP → Tau PET → structural MRI/clinical severity. The following thresholds were used for considering a good fit: CFI > 0.97; RMSEA < 0.05; SRMR < 0.05^30^. Associations were adjusted for age, sex, and (if not involving CDR-SB) cognitive status. For all analyses, we considered a two-sided P < 0.05 as statistically significant.

### 2.6 Proteomic co-expression network analysis

Weighted gene co expression network analysis (WGCNA) (v1.73) was used to identify modules of co-expressed proteins in the plasma proteomic dataset^31^. Soft-thresholding power was selected based on the scale-free topology criterion, with preference given to the lowest power at which the scale-free topology fit index approached an asymptote (typically *R*^2^ ≥ 0.90) while preserving acceptable network connectivity. Samples with more than 50% missing protein measurements were excluded, and proteins with more than 20% missing values across retained samples were removed before network construction. Modules were identified using the blockwiseModules () function with a minimum module size of 50 and a merge cut height of 0.15. Topological overlap matrices were computed from pairwise protein correlations to capture network interconnectedness. For each module, the module eigengene was defined as the first principal component of the scaled protein abundance matrix and used to represent the dominant expression pattern of that module. Furthermore, associations between module eigengenes and plasma GFAP levels were evaluated using linear regression models, with age, sex, and assay plate included as covariates. The reported β coefficients are standardized and represent the SD difference in module eigengene per 1-SD increase in plasma GFAP. Multiple comparisons were adjusted using the Benjamini–Hochberg procedure, and modules meeting an FDR *<* 0.05 were considered significantly associated with plasma GFAP. To assess whether the association with the yellow module was driven by the inclusion of GFAP itself, we performed a sensitivity analysis in which GFAP was excluded from the proteomic dataset and then repeated the WGCNA and the module–GFAP association analyses.

### 2.7 Functional enrichment analysis

To characterize the biological functions and signaling pathways associated with the plasma GFAP–related proteomic profile, we performed functional enrichment analyses on the 594 proteins comprised in the MEyellow module (M4) identified by WGCNA. Pathway enrichment analysis was performed using the enricher function from the cluster-Profiler (‘4.18.4’) R package. Gene sets were downloaded from MSigDB (v2025.1, human gene symbols, GMT format), including the Hallmark, Kyoto Encyclopedia of Genes and Genomes (KEGG), Reactome, Pathway Interaction Database (PID), ImmuneSigDB, and Human Phenotype Ontology (HPO). P values were corrected for multiple testing using the Benjamini–Hochberg method and pathways with an FDR-P corrected < 0.05 were considered significantly enriched.

### 2.8 Gene set variation analysis

To quantify pathway activity at the individual-sample level, gene set variation analysis (GSVA) was performed on the log-transformed protein level matrix using the GSVA R package (v2.4.4)^32^. The gene sets were the same as those used in the pathway enrichment analysis. The parameters: min.sz = 5, max.sz = 500 were set and other parameters were set to their default values. Single-sample gene set enrichment analysis (ssGSEA) scores were subsequently calculated for each pathway in each participant. Associations between ssGSEA scores and CSF Aβ42/40, Aβ PET, tau PET were assessed using linear regression models adjusted for age, sex, and assay plate. For this exploratory analysis, pathways were selected using P < 0.05 without multiple testing correction for each biomarker. Shared pathways were defined as those meeting this threshold for at least two biomarkers.

### 2.9 Protein–protein interaction network analysis

To investigate the interaction architecture of the hub proteins within GFAP-related pathways shared across CSF Aβ42/40, Aβ PET and tau PET, we constructed a protein–protein interaction (PPI) network using STRING (Version 12.0; https://string-db.org/)^33^. Proteins were mapped to STRING identifiers, and interactions for Homo sapiens were retrieved using a minimum confidence score of 0.7. Edges supported by all available evidence channels were retained, and isolated nodes were removed when appropriate. The network was visualized in Cytoscape (v3.10.4), and hub proteins were prioritized using maximal clique centrality implemented in the cytoHubba plugin (v0.1). Top-ranked proteins were considered candidate hub proteins within the GFAP-associated network.

## 3 Results

### 3.1 Participant Characteristics

The discovery cohort included 278 participants from the Knight ADRC who had plasma GFAP, CSF Aβ42/40, Aβ PET, tau PET, and structural MRI acquired within one year of the clinical assessment (age 69.4 ± 8.6 years, 51.8% female, 82.4% CN). The replication cohort comprised 236 participants from ADNI3 (ClinicalTrials.gov identifier: NCT02854033) with available plasma GFAP, plasma Aβ42/40, Aβ PET, tau PET, and structural MRI acquired within one year of the clinical assessment (age 71.6 ± 7.0 years, 55.5% female, 69.1% CN). Demographic and clinical characteristics are summarized in **Table 1**. Sample sizes and characteristics by A/T group (A−T−, A+T−, and A+T+) and for the subgroups (T− and A+) are provided in **Table S1** (Knight ADRC) and **Table S2** (ADNI3). The time intervals between biomarker measurements, imaging, and clinical assessments are reported for each cohort in **Table S3**. Soluble Aβ pathology was assessed using the CSF Aβ42/40 ratio in the discovery cohort and a plasma Aβ42/40 assay in the replication cohort, reflecting the best-available biofluid measure in each cohort. Compared with the discovery cohort, ADNI participants were significantly older (*P* = 0.019) and included a higher proportion of individuals with cognitive impairment (ADNI: 30.9% CDR ≥ 0.5; Knight ADRC: 17.6% CDR ≥ 0.5). In the discovery cohort, plasma proteomics (SomaScan 7K) data were available for 252 participants, with no significant differences in age, sex, APOE ε4 carrier status, or cognitive status between participants with and without plasma proteomics data (**Table S4**). An overview of the study design is depicted in **Figure S1**.

**Table 1.**
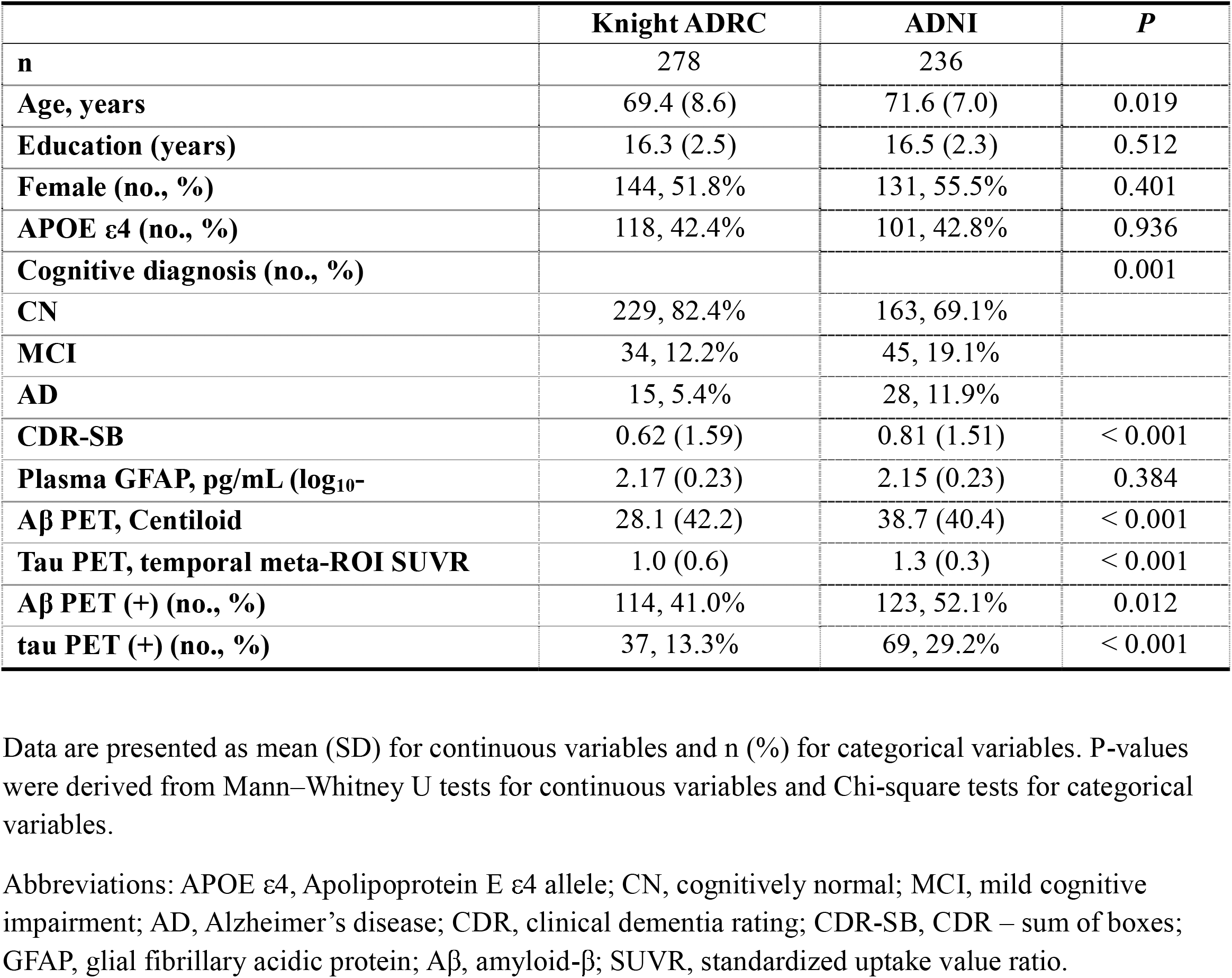
Participant characteristics by study cohort.

|  | **Knight ADRC** | **ADNI** | ***P*** |
|---|---|---|---|
| **n** | 278 | 236 |  |
| **Age, years** | 69.4 (8.6) | 71.6 (7.0) | 0.019 |
| **Education (years)** | 16.3 (2.5) | 16.5 (2.3) | 0.512 |
| **Female (no., %)** | 144, 51.8% | 131, 55.5% | 0.401 |
| **APOE ε4 (no., %)** | 118, 42.4% | 101, 42.8% | 0.936 |
| **Cognitive diagnosis (no., %)** |  |  | 0.001 |
| **CN** | 229, 82.4% | 163, 69.1% |  |
| **MCI** | 34, 12.2% | 45, 19.1% |  |
| **AD** | 15, 5.4% | 28, 11.9% |  |
| **CDR-SB** | 0.62 (1.59) | 0.81 (1.51) | < 0.001 |
| **Plasma GFAP, pg/mL (log<sub>10</sub>-** | 2.17 (0.23) | 2.15 (0.23) | 0.384 |
| **Aβ PET, Centiloid** | 28.1 (42.2) | 38.7 (40.4) | < 0.001 |
| **Tau PET, temporal meta-ROI SUVR** | 1.0 (0.6) | 1.3 (0.3) | < 0.001 |
| **Aβ PET (+) (no., %)** | 114, 41.0% | 123, 52.1% | 0.012 |
| **tau PET (+) (no., %)** | 37, 13.3% | 69, 29.2% | < 0.001 |

Data are presented as mean (SD) for continuous variables and n (%) for categorical variables. P-values were derived from Mann–Whitney U tests for continuous variables and Chi-square tests for categorical variables.
Abbreviations: APOE ε4, Apolipoprotein E ε4 allele; CN, cognitively normal; MCI, mild cognitive impairment; AD, Alzheimer’s disease; CDR, clinical dementia rating; CDR-SB, CDR – sum of boxes; GFAP, glial fibrillary acidic protein; Aβ, amyloid-β; SUVR, standardized uptake value ratio.

### 3.2 Associations between plasma GFAP and AD pathological biomarkers

In both cohorts, partial Spearman’s correlation analyses revealed that plasma GFAP levels were significantly associated with multiple core AD pathological biomarkers in the overall sample after adjustment for age, sex, and cognitive status, analyses involving CDR-SB were adjusted for age and sex only. To investigate these associations in subgroups defined by A/T status, participants were stratified into a tau PET-negative (T−) group (comprising A−T− and A+T− individuals) and an Aβ PET-positive (A+) group (comprising A+T− and A+T+ individuals). In T− individuals, plasma GFAP was associated with CSF Aβ42/40, Aβ PET, tau PET. In contrast, among A+ individuals, plasma GFAP was associated with Aβ PET, tau PET, structural MRI measures and clinical severity (**Figure 1a**). Similar patterns of association were observed in the replication cohort (**Figure 1f**).

**Figure 1.**
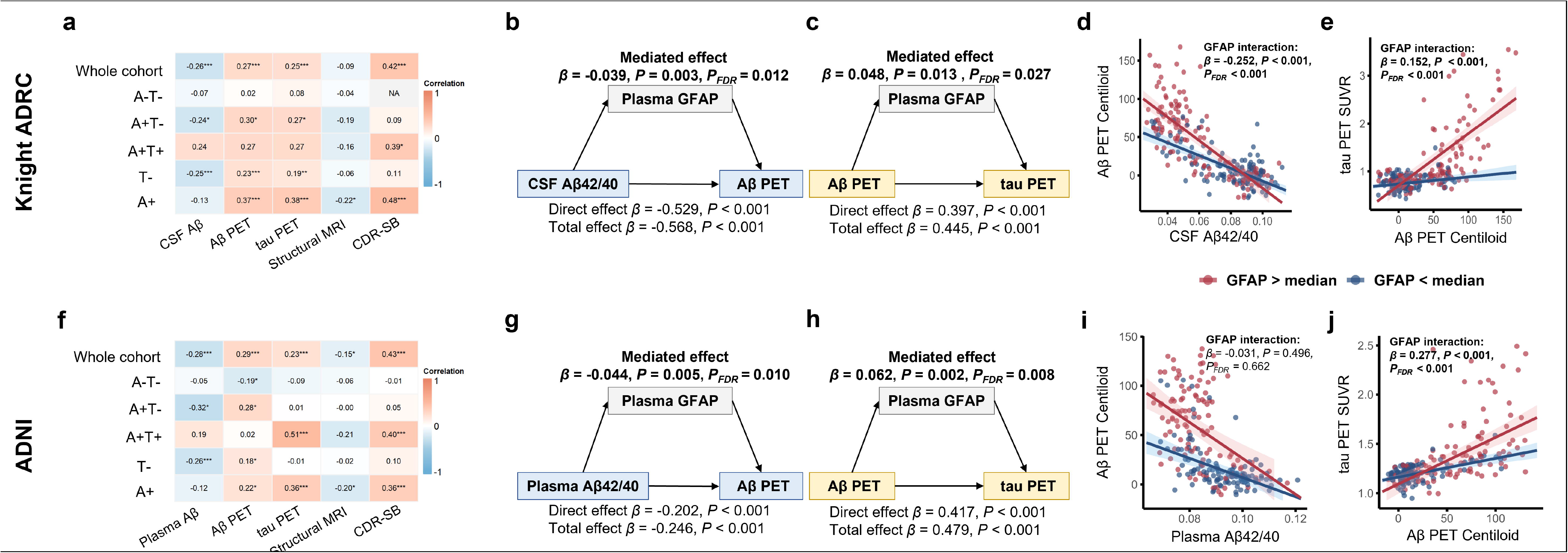
Plasma GFAP statistically mediates and moderates the associations between soluble Aβ, Aβ PET, tau PET, neurodegeneration and clinical severity across the AD continuum in the Knight ADRC (a-e, n=278) and ADNI (f-j, n=236) cohorts. a,f, Partial Spearman’s correlations illustrate the associations between plasma GFAP and core AD pathological biomarkers in the (a) Knight ADRC and (f) ADNI cohorts. b,g, Mediation analyses show that plasma GFAP partially mediates the associations between (b) CSF Aβ42/40 and Aβ PET burden in the Knight ADRC and (g) plasma Aβ42/40 and Aβ PET burden in the ADNI cohort. Solid paths denote statistically significant effects and dashed paths non–significant effects; standardized β coefficients are reported. c,h, Mediation analyses show that plasma GFAP also partially mediates the association between Aβ PET and tau PET in both cohorts. d,i, Scatter plots show that plasma GFAP significantly moderates the association between CSF Aβ42/40 and Aβ PET burden in the (d) Knight ADRC, but did not moderate the association between plasma Aβ42/40 and Aβ PET burden in the (i) ADNI cohort. e,j, Scatter plots show that plasma GFAP significantly moderates the association between Aβ PET and tau PET in both cohorts. β estimates and corresponding two-sided P values were computed from linear regression models. Shaded areas represent 95% confidence intervals of the regression lines. The P values for were adjusted using Benjamini–Hochberg false discovery rate (BH-FDR) correction, and the corrected P values are denoted as PFDR. Both unadjusted P and PFDR values are reported. NA, not estimable because CDR-SB scores had zero variance in the A−T− group in Knight ADRC.

### 3.3 Mediating and moderating roles of astrocytic reactivity in amyloid cascade across the AD continuum

We next systematically investigated the role of plasma GFAP across sequential steps of the AD pathological cascade (soluble Aβ → Aβ PET → tau PET → structural MRI/clinical severity). Mediation analyses showed that plasma GFAP partially mediated the associations between soluble Aβ and fibrillar Aβ deposition: CSF Aβ42/40 to Aβ PET in the Knight ADRC (indirect effect β = −0.039, 95% CI: −0.074 to −0.020, *P* = 0.003; **Figure 1b** and **Table S5**) and plasma Aβ42/40 to Aβ PET in the ADNI cohort (indirect effect β = −0.044, 95% CI: −0.082 to −0.019, *P* = 0.005; **Figure 1g** and **Table S5**). Plasma GFAP also significantly mediated the association between Aβ PET and tau PET in both cohorts (Knight ADRC: β = 0.048, 95% CI: 0.016 to 0.094, *P* = 0.013, **Figure 1c**; ADNI: β = 0.062, 95% CI: 0.029 to 0.111, *P* = 0.002, **Figure 1h**; Table S5).

Further, regression analyses revealed a significant interaction of CSF Aβ42/40 and plasma GFAP on Aβ PET in the Knight ADRC cohort (β = −0.252, 95% CI: −0.354 to −0.150, *P* < 0.001; **Figure 1d** and **Table S6**), whereas no such interaction was observed for plasma Aβ42/40 and plasma GFAP on Aβ PET in the ADNI cohort (β = −0.031, 95% CI: −0.122 to 0.059, *P* = 0.496; **Figure 1i** and **Table S6**). Plasma GFAP also significantly moderated the relationship between Aβ and tau PET in both cohorts (Knight ADRC: β = 0.152, 95% CI: 0.091 to 0.214, *P* < 0.001; **Figure 1e** and **Table S6**, ADNI: β = 0.277, 95% CI: 0.143 to 0.410, *P* < 0.001; **Figure 1j** and **Table S6**). All significant indirect effects and interaction terms remained significant after FDR correction (*PFDR* < 0.05; **Figure 1 and Table S5 and S6**). In contrast, GFAP showed no significant mediating or moderating effects on the downstream relationships between tau PET and structural MRI/clinical severity in either cohort (**Figure S2, Table S5 and S6**).

### 3.4 Mediating roles of astrocytic reactivity with amyloid and tau pathology within T− and A+ individuals

Before multiple-comparison correction, plasma GFAP significantly mediated the inverse association between soluble Aβ and Aβ PET burden in T− individuals across both cohorts (Knight ADRC: indirect effect β = −0.025, 95% CI: −0.051 to −0.008, *P* = 0.022; ADNI: β = −0.037, 95% CI: −0.087 to −0.009, *P* = 0.047). After FDR correction, the effect remained significant only in Knight ADRC (*Pfdr* = 0.044), not in ADNI (*Pfdr* = 0.094; **Figure 2a, g and Table S7**). No mediating effect of plasma GFAP was observed for the association between Aβ PET and tau PET (**Figure 2b, h** and **Table S7**). In contrast, in A+ individuals, plasma GFAP mediated the association between Aβ PET and tau PET across both cohorts (Knight ADRC: indirect effect β = 0.070, 95% CI: 0.012 to 0.156, *P* = 0.045; ADNI: β = 0.065, 95% CI: 0.021 to 0.134, *P* = 0.021), but the indirect effect remained significant after FDR correction only in ADNI **(*PFDR* = 0.042; Figure 2d, j and Table S7**). No significant indirect effect through plasma GFAP was observed between soluble Aβ42/40 and Aβ PET in either cohort (**Figure 2c, i and Table S7**).

**Figure 2.**
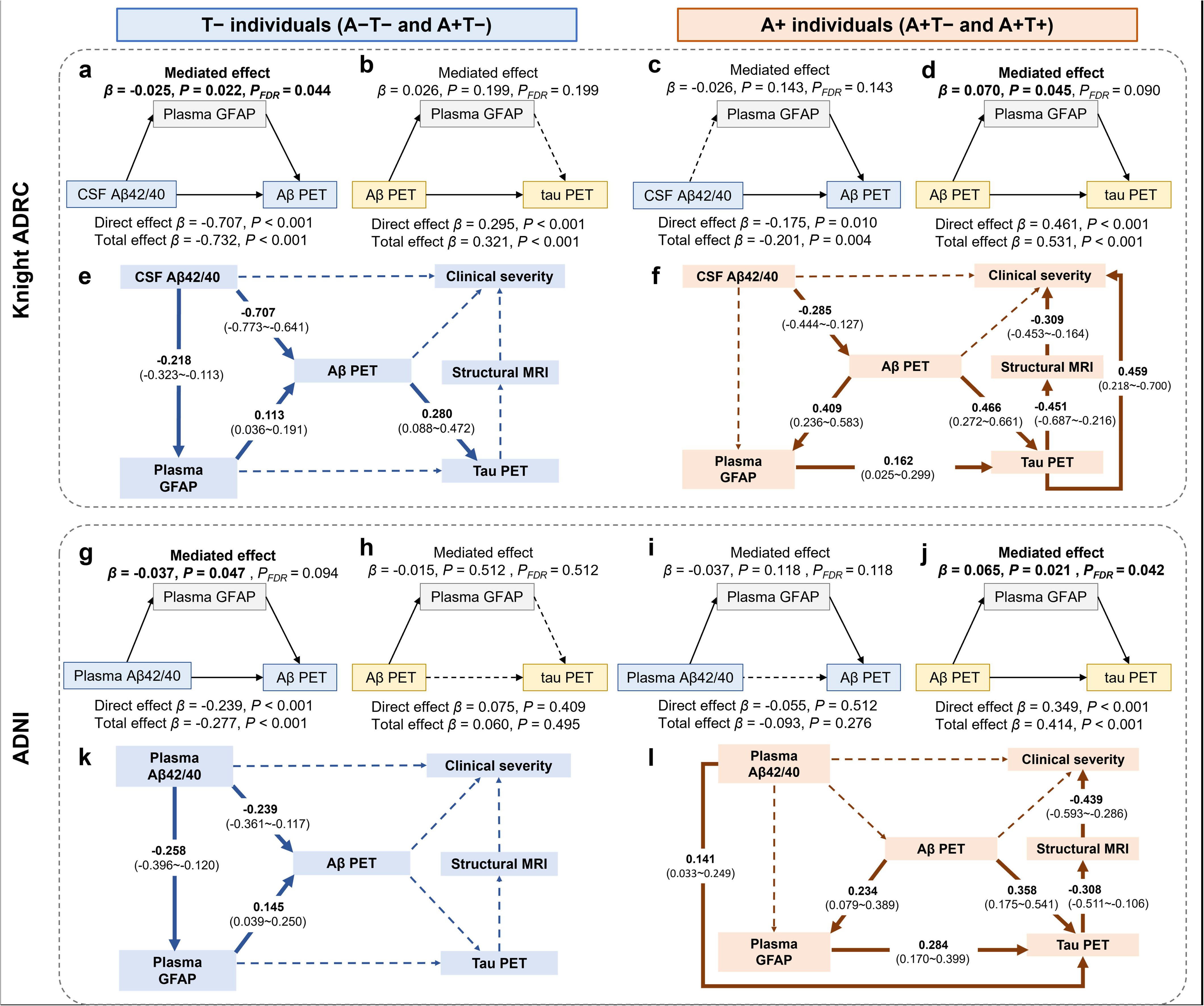
Mediating role of plasma GFAP with amyloid and tau pathology within T− and A+ individuals. a,b,g,h, In T− individuals, plasma GFAP statistically mediates the associations between (a) CSF Aβ42/40 and Aβ PET burden in the Knight ADRC and (g) plasma Aβ42/40 and Aβ PET burden in the ADNI cohort, but this effect did not survive FDR correction with plasma Aβ42/40 in ADNI. In contrast, plasma GFAP showed no significant mediating effect on the association between Aβ PET and tau PET in either cohort (b,h). c,d,i,j, In A+ individuals, plasma GFAP significantly mediates the association between Aβ PET and tau PET in both the (d) Knight ADRC and (j) ADNI cohorts, but survived correction only in ADNI. In contrast, plasma GFAP showed no significant mediating effect on the association between soluble Aβ markers and Aβ PET burden (c,i). Solid paths denote statistically significant effects and dashed paths non–significant effects; standardized β coefficients are reported. e,k, In T− individuals, structural equation modeling characterizing the associations among soluble Aβ (CSF Aβ42/40 or plasma Aβ42/40), plasma GFAP, Aβ PET, tau PET, structural MRI, and clinical severity in the (e) Knight ADRC and (k) ADNI cohorts. f,l, In A+ individuals, structural equation modeling testing the associations among soluble Aβ (CSF Aβ42/40 or plasma Aβ42/40), plasma GFAP, Aβ PET, tau PET, structural MRI, and clinical severity in the (f) Knight ADRC and (l) ADNI cohorts. Solid lines with standardized β estimates and 95% CI represent significant associations, and dashed lines represent non-significant effects. All analyses were adjusted for age, sex, and cognitive status, except those involving CDR-SB, which were adjusted for age and sex only. The P values for were adjusted using Benjamini–Hochberg false discovery rate (BH-FDR) correction, and the corrected P values are denoted as PFDR. Both unadjusted P and PFDR values are reported.

We further applied SEM to test the associations among plasma GFAP, soluble Aβ, Aβ PET, tau PET, structural MRI and clinical severity according to A/T biomarker status. In T− individuals, plasma GFAP mediated the association between soluble Aβ (CSF Aβ42/40 or plasma Aβ42/40) and Aβ PET, with no significant pathways to tau PET, structural MRI, or clinical severity in both the Knight ADRC (**Figure 2e**) and ADNI (**Figure 2k**) cohorts. Conversely, in A+ individuals, GFAP mediated the association between Aβ PET and tau PET, which was further associated with clinical severity through tau aggregation and brain atrophy, consistently observed in both the Knight ADRC (**Figure 2f**) and ADNI (**Figure 2l**) cohorts. The SEM in T− individuals from Knight ADRC showed mixed fit indices (X^2^ = 3.573, degrees of freedom = 1, RMSEA = 0.097, SRMR = 0.007, CFI = 0.997). The remaining models fit the data well (X^2^ = 0.119∼1.066, degrees of freedom = 1, RMSEA = 0.000∼0.009, SRMR = 0.002∼0.007, CFI = 1). **Tables S8 and S9** report detailed coefficients and associated statistics for structural equation models.

### 3.5 Moderating roles of astrocytic reactivity with amyloid and tau pathology within T− and A+ individuals

The subgroup interaction findings were not consistently replicated across cohorts. In T− individuals, a significant interaction between CSF Aβ42/40 and plasma GFAP on Aβ PET was observed in the Knight ADRC cohort (β = −0.180, 95% CI: −0.261 to −0.100, *P* < 0.001, *PFDR* < 0.001; **Figure 3a** and **Table S10**). As illustrated in **Figure 3e** (left), this interaction effect was primarily localized to classical AD-vulnerable cortical regions. Stratified analyses in T− individuals indicated that the association between CSF Aβ42/40 and Aβ PET was stronger in the high-GFAP group (β = −0.873, 95% CI: −0.999 to −0.748, *P* < 0.001) than in the low-GFAP group (β = −0.523, 95% CI: −0.636 to −0.410, *P* < 0.001; **Table S10**). In region-wise linear regressions, we observed that lower CSF Aβ42/40 levels were more strongly associated with higher Aβ PET burden mainly in neocortical brain regions in the high-GFAP group (**Figure 3g**), suggesting a globally amplified effect of astrocytic reactivity on amyloid accumulation. In contrast, no significant interaction was observed for plasma Aβ42/40 in the ADNI cohort (**Figure S3a** and **Table S10**). Additionally, no significant interaction between Aβ PET and plasma GFAP on tau PET was observed in T− individuals in either the Knight ADRC cohort (**Figure 3b**) or the ADNI cohort (**Figure S3b**).

**Figure 3.**
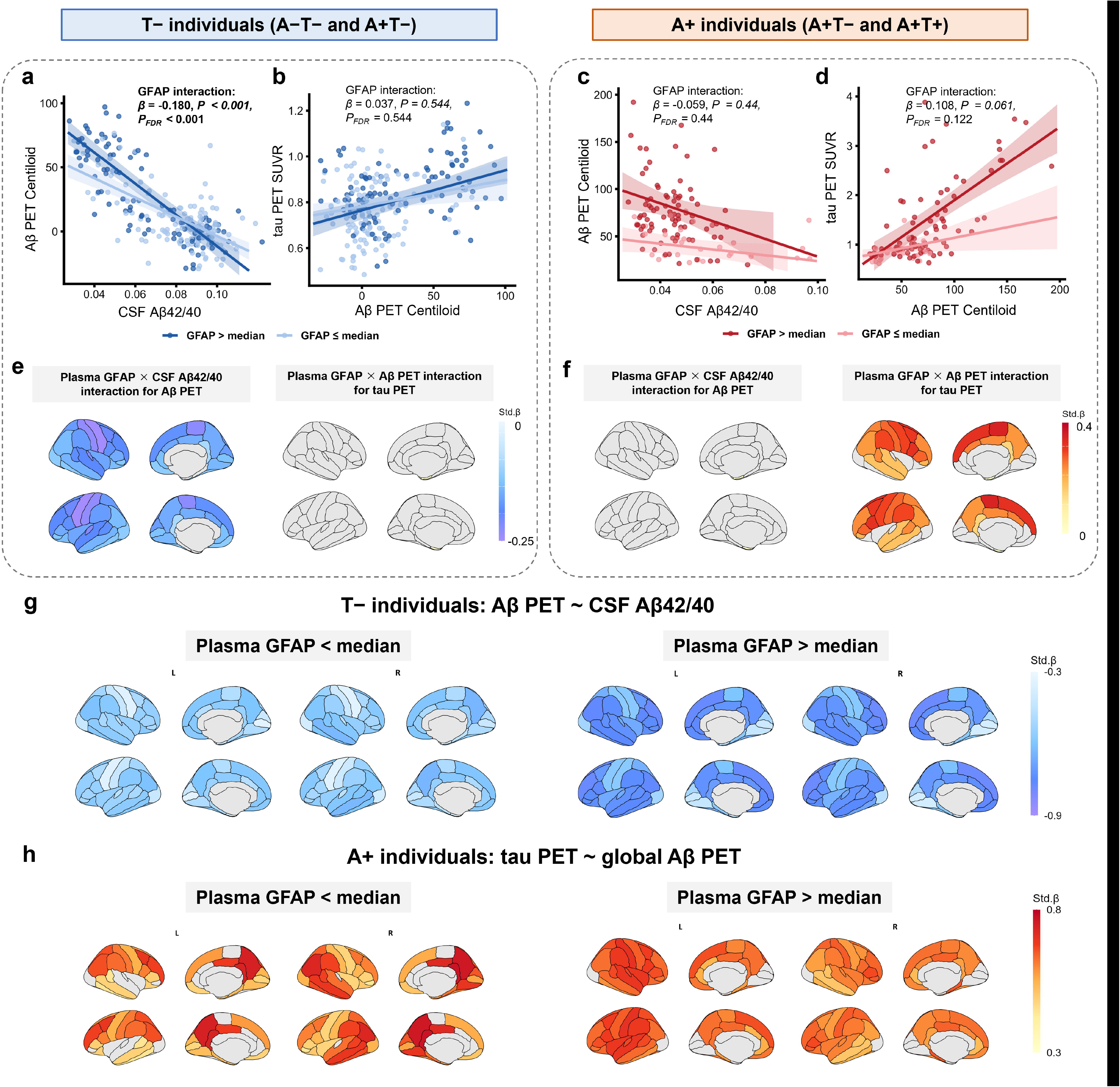
Moderating role of plasma GFAP with amyloid and tau pathology in the Knight ADRC cohort within T− and A+ individuals. a-d, Scatter plots displaying the association between CSF Aβ42/40 and Aβ PET burden, and between Aβ PET and tau PET according to plasma GFAP status in (a,b) T− and (c,d) A+ individuals. β estimates and corresponding two-sided P values were computed from linear regression models. Shaded areas represent 95% confidence intervals of the regression lines. e,f, Region-wise linear regression βmap (FDR-corrected P < 0.05) showing the interaction effects between CSF Aβ42/40 and plasma GFAP on Aβ PET burden, and between Aβ PET burden and plasma GFAP on tau PET across cortical regions in (e) T− and (f) A+ individuals. Regression models with standardized biomarker levels were adjusted for age, sex, and cognitive status, as well as the main effects of each interacting variables. g, In T− individuals, region-wise linear regression βmap (FDR-corrected P < 0.05) showing the association between CSF Aβ42/40 and Aβ PET burden across cortical areas in individuals in the low–GFAP group (left) and high–GFAP group (right). h, In A+ individuals, region-wise linear regression βmap (FDR-corrected P < 0.05) showing the association between Aβ PET burden and tau PET across cortical areas in low–GFAP group (left) and high–GFAP group (right) individuals. Regression models with standardized biomarker levels were adjusted for age, sex, and cognitive status. The P values for were adjusted using Benjamini– Hochberg false discovery rate (BH-FDR) correction, and the corrected P values are denoted as PFDR. Both unadjusted P and PFDR values are reported.

In A+ individuals, plasma GFAP did not significantly moderate the association between soluble Aβ (CSF/plasma Aβ42/40) and Aβ PET in either Knight ADRC (**Figure 3c**) or ADNI cohort (**Figure S3c**). In contrast, the interaction effect between plasma GFAP and Aβ PET on tau PET in A+ individuals exhibited a trend in the Knight ADRC cohort (β = 0.108, 95% CI: −0.005 to 0.221, *P* = 0.061, *PFDR* = 0.122; **Figure 3d** and **Table S10**), whereas it reached statistical significance in the ADNI cohort (β = 0.363, 95% CI: 0.079 to 0.647, *P* = 0.013, *PFDR* = 0.026; **Figure S3d** and **Table S10**). Although the interaction with global amyloid burden did not reach statistical significance in the discovery cohort, region-wise analyses showed that the interaction effect was significantly localized to AD-vulnerable regions, including the posterior cingulate cortex, precuneus, lateral parietal, and temporal association cortices (**Figure 3f** [right]). Stratified analyses in A+ individuals further demonstrated that the association between Aβ PET and tau PET was stronger in the high– compared to the low–GFAP group (**Table S10**), with stronger and more spatially widespread Aβ–tau coupling at higher GFAP levels (**Figure 3h**).

In an exploratory sensitivity analysis restricted to T− individuals with Aβ PET < 50 Centiloids, the plasma GFAP × CSF Aβ42/40 interaction on Aβ PET remained significant (β = −0.150, *P* = 0.015, **Figure S4c**), and the inverse association between CSF Aβ42/40 and Aβ PET was stronger in the high-GFAP group than in the low-GFAP group. The interaction was also significant at thresholds of 52, 54, and 56 Centiloids (*P* < 0.05), but not at 46 and 48 Centiloids (**Figure S4**).

### 3.6 Protein co-expression network analysis reveals protein modules associated with plasma GFAP

To further characterize proteomic signatures of astrocytic reactivity, WGCNA was applied to the plasma proteomic (SomaScan 7K) data from a subset of Knight ADRC participants (n = 252; **Table S4**). This analysis identified 7 distinct protein co-expression modules (module sizes: 100– 1,843 proteins). Among these, the MEyellow module (M4) emerged as the only module significantly associated with plasma GFAP (β = 0.25, *PFDR* < 0.001, **Figure 4a, Table S11**). After excluding GFAP and repeating the WGCNA, the re-derived yellow module remained significantly associated with plasma GFAP (β = 0.25, *PFDR* = 0.001; Figure S5). Functional annotation of 594 proteins within this module (**Table S12**) revealed enrichment in biological processes related to cell recognition, neuron development, and glycosaminoglycan binding.

**Figure 4.**
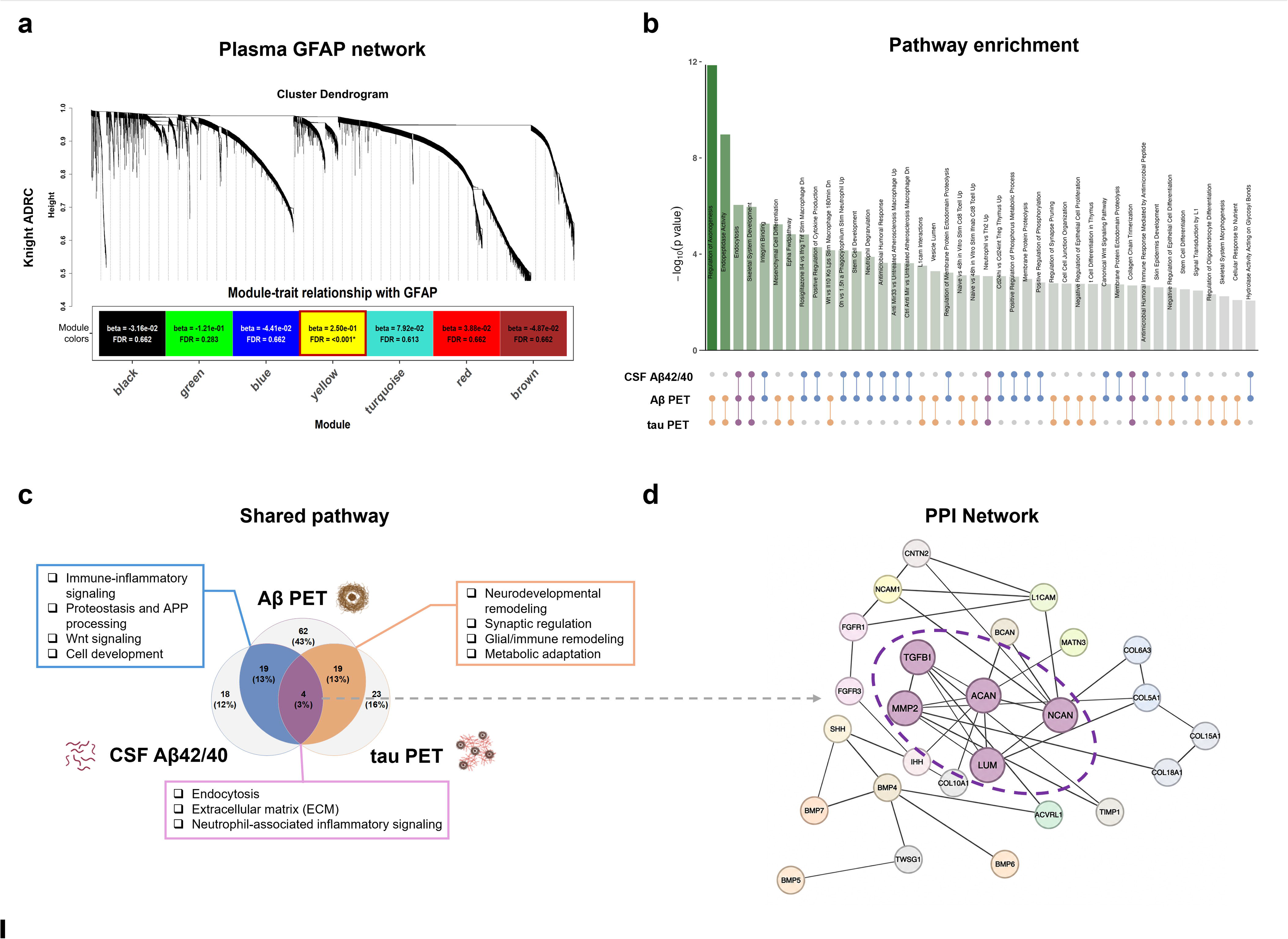
Proteomic profiling identifies plasma GFAP-associated pathways and hub proteins linking soluble Aβ, Aβ deposition and tau aggregation. a, Cluster dendrogram of plasma proteomic profiles generated by weighted gene co-expression network analysis (WGCNA), with proteins grouped into distinct co-expression modules. Heatmap showing correlations between module eigengenes and plasma GFAP. The yellow module (M4) shows a significant association with plasma GFAP after false discovery rate (FDR) correction and was selected for downstream analyses. β represents the SD change in module eigengene per 1-SD increase in plasma GFAP. b, Pathway enrichment analysis of the GFAP associated yellow module identified by WGCNA. Dot chart illustrating several pathways shared across CSF Aβ42/40, Aβ PET, and tau PET. c, The Venn diagram shows the overlap of identified pathways associated with CSF Aβ42/40, Aβ PET, and tau PET based on gene set variation analysis (GSVA) scores, with unadjusted P < 0.05 used for pathway selection. Shared pathways between soluble Aβ and Aβ PET are shown in blue, shared pathways between Aβ PET and tau PET are shown in orange, and pathways commonly shared across all three biomarkers are depicted in purple. d, Protein–protein interaction (PPI) network analysis based on pathways shared across CSF Aβ42/40, Aβ PET burden, and tau PET, identifying TGFB1, ACAN, NCAN, MMP2, and LUM as central hub proteins within an astrocyte-centered extracellular matrix–inflammatory network. Aβ, amyloid beta; CSF, cerebrospinal fluid; ECM, extracellular matrix; APP, amyloid precursor protein.

Pathway analysis further revealed significant enrichment of immune-related pathways, extracellular matrix (ECM) organization, Wnt signaling, and growth factor binding (**Figure S6**), suggesting a coordinated network of inflammatory processes, ECM remodeling and neurodevelopmental signaling.

### 3.7 Identification of key pathways and proteins associated with soluble Aβ, Aβ deposition and tau aggregation

To further quantify pathway activity at the individual-sample level, GSVA was performed for the significantly enriched pathways identified from the MEyellow module (M4). We then examined the associations between pathway activity and the three AD pathological biomarkers, including CSF Aβ42/40, Aβ PET and tau PET. These exploratory associations were evaluated at an unadjusted P < 0.05. Pathways associated with CSF Aβ42/40 were predominantly enriched for immune-inflammatory activation, ECM remodeling, and growth factor binding, consistent with astrocyte-centered inflammatory and matrix responses (**Table S13**). In contrast, pathways associated with Aβ PET additionally involved neurodevelopmental signaling and synaptic/neuronal regulation, suggesting broader circuit-level remodeling accompanying fibrillar amyloid accumulation (**Table S14**). Tau PET–associated pathways were primarily enriched for synaptic/neuronal regulation, ECM remodeling, and developmental morphogenesis (**Table S15**).

Specifically, pathways shared between CSF Aβ42/40 and Aβ PET were enriched for immune-inflammatory signaling, endocytosis, proteolysis and amyloid precursor protein (APP) processing, Wnt signaling, and stem-cell-related processes (**Figure 4c** and **Table S16**).

Conversely, pathways jointly associated with Aβ PET burden and tau PET were enriched for neurodevelopmental remodeling, synaptic regulation, cell adhesion, glial/immune responses, and metabolic adaptation (**Figure 4c** and **Table S17**). No pathways were uniquely shared between CSF Aβ42/40 and tau PET. Notably, endocytosis, ECM remodeling, and neutrophil-associated inflammatory signaling emerged as convergent pathways shared across CSF Aβ42/40, Aβ PET, and tau PET (**Figure 4c** and **Table S18**). Finally, to identify key proteins acting as common bridges linking plasma GFAP to soluble Aβ, Aβ PET, and tau PET, we performed PPI network analysis on proteins involved in the convergent pathways. This analysis identified TGFB1, ACAN, NCAN, MMP2, and LUM as hub proteins in the shared network (**Figure 4d**). These proteins are functionally involved in extracellular matrix organization (aggrecan [ACAN], neurocan [NCAN], and lumican [LUM]), perineuronal and matrix remodeling (matrix metalloproteinase 2 [MMP2] and NCAN), and inflammatory signaling (transforming growth factor beta 1 [TGFB1] and MMP2), suggesting a coordinated astrocytic-centered ECM–immune remodeling program across the AD continuum.

## 4 Discussion

In this study, using a discovery cohort and an independent replication cohort, we found that plasma GFAP mediated the association of soluble Aβ42/40 with Aβ PET, as well as the association of Aβ PET with tau PET in the overall samples, which remained significant after false discovery rate (FDR) correction. In the absence of tau pathology, plasma GFAP mediated the association between soluble Aβ alterations and fibrillar Aβ deposition. In stages characterized by established Aβ pathology, plasma GFAP mediated the association between Aβ PET and tau PET. These effects were similar in direction across cohorts, but were only statistically significant in one cohort after FDR correction. In our evaluation of the associations between GFAP and proteomics data, plasma GFAP–related astrocytic involvement in the amyloid cascade converges on an astrocyte-centered ECM–inflammatory axis. This may inform future diagnostic panels or targeted therapies.

In the overall sample, plasma GFAP mediated the relationships between soluble Aβ42/40 and Aβ deposition, and between Aβ and tau deposition, but not between tau pathology and structural MRI measures or clinical severity. This aligns with current evidence that plasma GFAP selectively engages amyloid-driven processes more strongly than downstream neurodegenerative changes^3^. Specifically, in individuals without tau pathology, plasma GFAP both mediated and moderated the relationship between soluble amyloid dysregulation (CSF Aβ42/40) to fibrillar amyloid deposition (Aβ PET burden). This finding suggests a key role for astrogliosis in early Alzheimer’s disease pathogenesis, aligning with recent research suggesting that astrocytic responses in the “conversion” from soluble Aβ changes to detectable plaque burden^34^. Consistent with this, SEM in a preclinical cohort found an inverse bidirectional relationship between CSF Aβ42/40 and plasma GFAP, supporting GFAP as a biomarker of astrocytic reactivity associated with early Aβ dysmetabolism^35^. However, these cross-sectional findings do not establish the temporal or causal direction of this relationship. Our finding that GFAP mediates the association between soluble Aβ42/40 (CSF in the Knight ADRC; plasma in ADNI) and Aβ burden should be interpreted as an indirect association and does not exclude alternative or reciprocal directions.

The interaction observed with CSF Aβ42/40 compared with plasma Aβ42/40 may further suggest that astrocytic responses are more tightly coupled to central amyloid dynamics than to peripheral measures. Importantly, our sensitivity analysis restricted to T− individuals with Aβ PET < 50 Centiloids still showed a robust GFAP × CSF Aβ42/40 interaction, supporting that GFAP-indexed astrocytosis may modify amyloid-related associations even at lower amyloid burden, before detectable tau pathology. In contrast, at stages characterized by Aβ positive, GFAP mediated the association between Aβ PET and tau PET, which was further associated with brain atrophy and clinical severity. These results extend prior work showing that plasma GFAP partially explained the effect of Aβ PET on tau PET or CSF/ plasma p-tau, and linked Aβ pathology to subsequent neurodegeneration and cognitive decline^36^. This supports the notion that tau burden and its propagation are more tightly linked to neurodegeneration and cognitive decline.

Although the stratified mediation findings were consistent in direction across cohorts, not all were significant following FDR correction. The interaction of CSF Aβ42/40 × plasma GFAP on Aβ PET was observed only in the Knight ADRC cohort and did not replicate with plasma Aβ42/40 in ADNI, possibly reflecting differences in the available biofluid biomarkers across cohorts rather than biological inconsistency. Furthermore, in the Aβ-positive individuals, the interaction between Aβ PET × plasma GFAP on tau PET reached statistical significance only in the ADNI cohort, whereas a similar trend was observed in the Knight ADRC cohort, plausibly reflecting differences in disease stage composition, as ADNI contains a greater proportion of individuals at later stages of AD severity. However, these possible explanations remain speculative. Region-wise analyses in the relatively younger and less cognitively impaired Knight ADRC cohort localized significant interaction effects to parietal and temporal regions, suggesting that Aβ-related astrocytic reactivity may enhance tau accumulation particularly in AD-vulnerable cortical regions. Accordingly, the moderation findings should be considered hypothesis-generating rather than validated, and future studies using harmonized biomarker assays are needed to resolve these discrepancies.

These findings help reconcile apparently conflicting evidence on how GFAP relates to AD pathology. In preclinical or cognitively unimpaired cohorts, plasma GFAP has consistently emerged as an early marker sensitive to Aβ pathology^3,34^. Studies conducted in clinical and memory-clinic cohorts more often report associations between higher GFAP and greater tau tangle burden, cortical thinning, and faster cognitive decline, suggesting that serum GFAP is more strongly associated with tau tangles than amyloid plaques in later advanced AD stages^37,38^. Together, these findings align with a proposed “two-wave” model of astrocytic reactivity, with an initial Aβ-related wave in presymptomatic or prodromal stages, and a second wave at more advanced stages of Alzheimer’s disease dementia^39^. In this framework, our findings suggest that astrocytic reactivity was associated with the relationship between soluble Aβ and fibrillar Aβ deposition in T− individuals and between Aβ and tau accumulation in A+ individuals.

Proteomic analyses in the Knight ADRC cohort provide an integrated view of the biological basis underlying astrocytic reactivity across the AD continuum, aligning with the evidence that reactive astrocytes exist in multiple functionally distinct sub-states rather than a unitary state^40,41^. The GFAP-associated protein module was enriched in immune, ECM organization, wnt signaling and growth factor pathways, providing cross-modal support for experimental and transcriptomic evidence that reactive astrocytes upregulate proteostasis and inflammatory programs in response to misfolded proteins such as Aβ^42–44^. Shared pathways between CSF Aβ42/40 and Aβ PET included immune-inflammatory signaling, APP processing, endocytosis, and Wnt-related pathways, suggesting that amyloid deposition unfolds within a coordinated immune-reactive and proteostasis-related milieu^45,46^. In contrast, pathways overlapping Aβ PET and tau PET highlighted synaptic regulation, neurodevelopmental remodeling, glial/immune activation, and metabolic adaptation, echoing reports that tau–linked astrocyte states are tightly coupled to synaptic, neuronal and network dysfunction underlying neurodegeneration and cognitive decline.^47^ The convergence of endocytosis, ECM remodeling, and neutrophil-associated inflammatory signaling across the soluble Aβ, fibrillar Aβ, and tau pathology implicates a core protein–protein interaction hub centered on TGFB1, ACAN, NCAN, MMP2 and LUM, pointing to an astrocyte-centered ECM–inflammatory axis that may link amyloid and tau pathology^42^. Consistent with the “two-wave” model of astrocytic reactivity in AD, this axis positions astrocytes as neuroimmunocytes that orchestrate both inflammatory signaling and extracellular remodeling. Crucially, plasma GFAP–related astrocytic involvement in amyloid cascade converges on an ECM–inflammatory axis, highlighting this pathway as a potential target for future therapeutic strategies.

The strengths of our study include a relatively large, well–characterized sample, the integration of fluid biomarkers with multimodal imaging data (including in vivo quantification of Aβ, tau, and neurodegeneration), and the availability of plasma proteomics data in the discovery cohort. Several limitations should also be acknowledged. First, Aβ pathology was assessed using different biomarkers across cohorts (CSF Aβ42/40 in the Knight ADRC; plasma Aβ42/40 in the ADNI cohort), which limits direct comparison between cohorts. Second, the cross-sectional design of the present study limits causal inferences. The directionality specified in our mediation models reflects the hypotheses under investigation rather than an established biological sequence. Third, after FDR correction, the mediation effects stratified by A/T stages remained significant in only one cohort each, whereas the other cohort showed an effect of similar direction and magnitude. These stage-specific associations should therefore be considered preliminary and warrants replication in independent, longitudinal cohorts. Lastly, to better understand the neuroinflammatory response in AD, future research should also explore other astroglia biomarkers, such as YKL-40 and S100B, as well as microglial markers like sTREM2 in conjunction with GFAP. Future longitudinal studies and experimental models are needed to validate these mechanistic insights and translate them into clinical applications.

In conclusion, our findings suggest that plasma GFAP is associated with the relationship between soluble Aβ42/40 and Aβ PET in tau-negative individuals, and with the relationship between Aβ PET and tau PET in Aβ-positive individuals. Exploratory plasma proteomics further identify an ECM–inflammatory axis as a promising candidate for targeted intervention in AD.

## Data Availability

Data from the Knight ADRC can be requested by qualified investigators (https://knightadrc.wustl.edu/professionals-clinicians/request-center-resources/). Data from the ADNI can be requested via the LONI website (https://adni.loni.usc.edu/).

## ACKNOWLEDGMENTS

The authors thank all research volunteers for their contributions.

## AUTHOR CONTRIBUTIONS

X.N. and Q.W. designed and supervised the study. X.N. and Q.S. processed and analyzed the data. X.N., Q.S., and Q.W. interpreted the data. J.W., M.J., A.L., D.A.H., S.E.S., C.C., B.A.G., Y.W., and T.L.S.B. contributed to data acquisition, data interpretation, and scientific discussion. X.N. wrote the manuscript. All authors reviewed, edited, and approved the final manuscript.

## CONSENT STATEMENT

All study participants provided written informed consents for all study procedures.

## SUPPORTING INFORMATION

Additional supporting information can be found online in the Supporting Information section at the end of this article.

