## Supplementary figures and tables for "Stage–Specific Roles of Plasma GFAP in Linking Amyloid Cascade in Alzheimer Disease": Supplementary material.docx

**Supplementary Methods |** Structural MRI

**Figure S1 |** Study design and workflow.

**Figure S2 |** Plasma GFAP does not significantly mediate or moderate the associations between tau pathology and neurodegeneration/clinical severity.

**Figure S3 |** Moderating roles of astrocytic reactivity with amyloid and tau pathology in A/T-defined subgroups of the ADNI cohort.

**Figure S4 |** Sensitivity analyses: plasma GFAP × CSF Aβ42/40 interaction on Aβ PET in tau-negative individuals with relatively lower amyloid burden across Centiloid thresholds.

**Figure S5 |** Sensitivity analysis excluding GFAP from the WGCNA input.

**Figure S6 |** Functional enrichment analyses of proteins in the MEyellow module.

**Table S1 |** Participant characteristics by A/T status in Knight ADRC cohort.

**Table S2 |** Participant characteristics by A/T status in ADNI cohort.

**Table S3 |** Time differences between imaging, fluid, and clinical assessments in the Knight ADRC and ADNI cohorts.

**Table S4 |** Participant characteristics of the Knight ADRC cohort according to availability of plasma proteomics data.

**Table S5 |** Mediation effects of plasma GFAP for the association between different components of the AD cascade.

**Table S6|** Moderation effects of plasma GFAP for the association between different components of the AD cascade.

**Table S7 |** Mediating roles of astrocytic reactivity with amyloid and tau pathology within T− and A+ individuals.

**Table S8 |** Structural equation model coefficients associated statistics for Fig. 2 e and k. Reported are standardized β (95% CI) P-value and FDR-adjusted p-values.

**Table S9 |** Structural equation model coefficients associated statistics for Fig. 2 f and l. Reported are standardized β (95% CI) P-value and FDR-adjusted p-values.

**Table S10 |** Moderating roles of astrocytic reactivity with amyloid and tau pathology within T− and A+ individuals.

**Table S11 |** Associations between plasma GFAP and protein modules.

Abbreviations: AD, Alzheimer’s disease; APOE ε4, Apolipoprotein ε4 allele; CN, cognitively normal; MCI mild cognitive impairment; CDR, clinical dementia rating; CDR-SB, CDR – sum of boxes; MMSE, mini-mental state examination; GFAP, glial fibrillary acidic protein; Aβ, amyloid-β; SUVR, standardized uptake value ratio; CI, confidence interval.

Supplementary Methods

**Structural MRI**

Imaging was performed using either a 3.0 Tesla Siemens Biograph mMR (Erlangen, Germany) or 3.0 Tesla Siemens TIM Trio (Erlangen, Germany) scanner. For the Siemens Biograph mMR, high-resolution 3-dimensional sagittal T1 magnetization prepared – rapid gradient echo (MP-RAGE) anatomical images were acquired with the scanning parameters of repetition time (TR) = 2300 ms, time to echo (TE) = 2.95 ms, flip angle = 9°, 176 slices, acquisition matrix = 240 x 256, and voxel size = 1 x 1 x 1.2 mm^3^. For the Siemens TIM Trio, high-resolution 3-dimensional sagittal T1 MP-RAGE anatomical images were acquired with the scanning parameters of TR = 2400 ms, TE = 3.16 ms, flip angle = 8°, 176 slices, acquisition matrix = 256 x 256, and voxel size = 1 x 1 x 1 mm^3^.

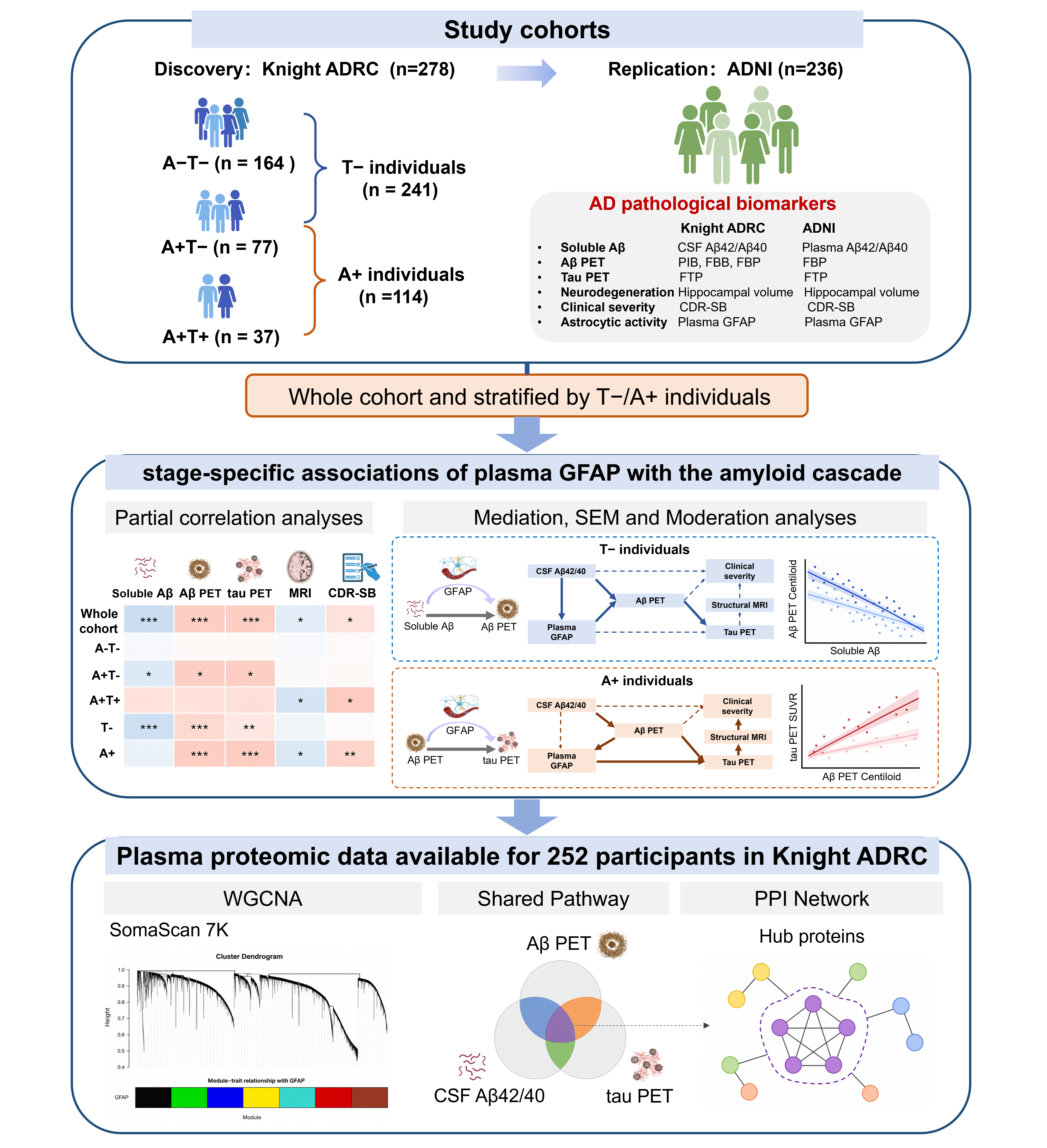

**Figure S1** **Study design and workflow.**

Participants from the Knight ADRC (discovery) and ADNI (replication) cohorts with available Alzheimer’s disease (AD) pathological biomarkers, including soluble Aβ measures (CSF Aβ42/40 in Knight ADRC and plasma Aβ42/40 in ADNI), Aβ PET, tau PET, structural MRI, clinical severity (CDR-SB) were included in the analysis. Participants were stratified into A−T−, A+T−, and A+T+ groups, and further grouped into T− (A−T− and A+T−) and A+ (A+T− and A+T+) subgroups. First, we investigated the role of astrocytic reactivity in the AD pathological cascade using partial correlation, mediation, moderation, and SEM analyses in both cohorts. Second, to characterizing the biological basis underlying astrocytic reactivity in AD, we analyzed plasma proteomic data from a subset of Knight ADRC participants (n = 252). Plasma samples were analyzed using the SomaScan assay, measuring a total of 6,905 analytes targeting 6,106 proteins. WGCNA, pathway enrichment analysis and protein–protein interaction (PPI) network analysis were performed to identify GFAP-associated pathways and hub proteins linked to soluble Aβ dysregulation, Aβ and tau deposition. Aβ, amyloid-β; GFAP, glial fibrillary acidic protein; PET, positron emission tomography; PPI, protein–protein interaction; SEM, structural equation modeling; WGCNA, weighted gene co-expression network analysis.

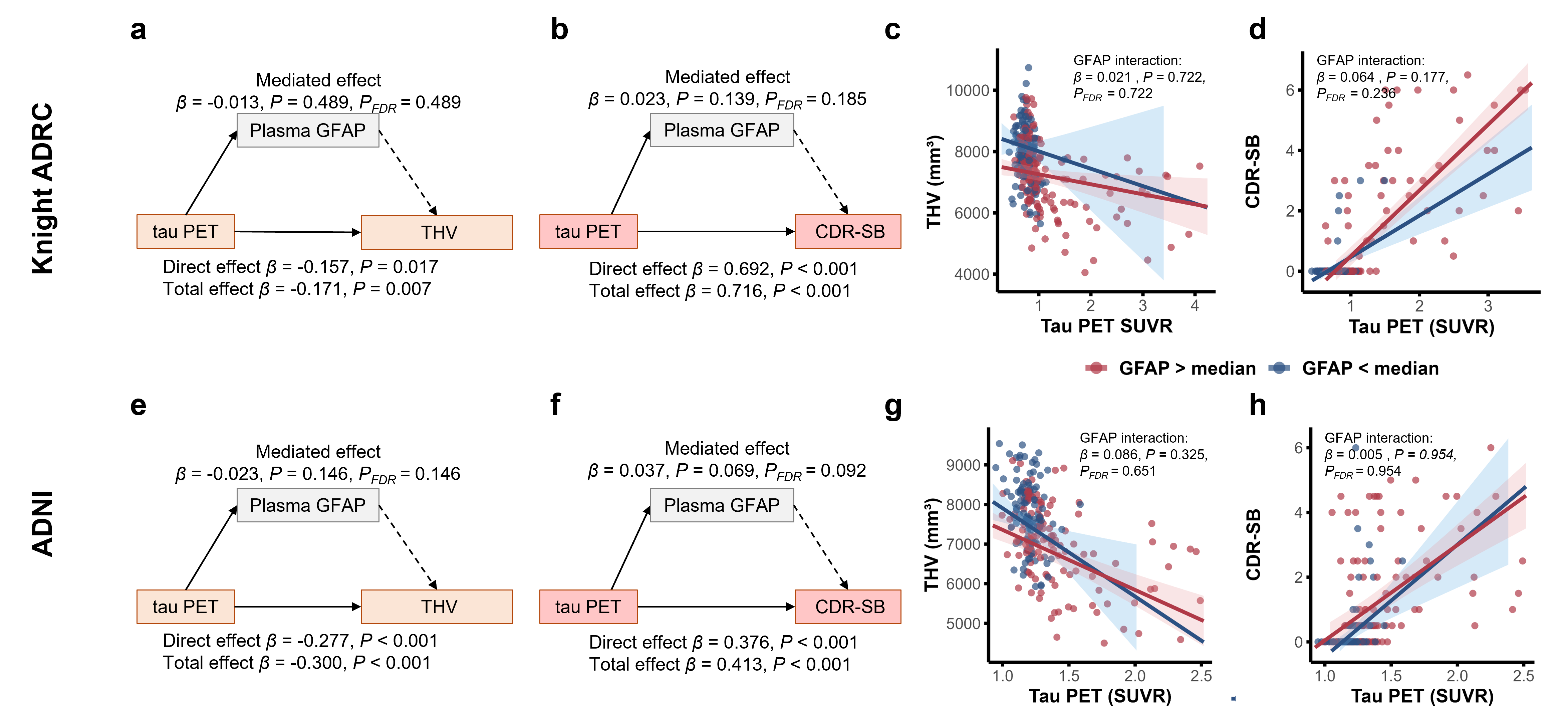

**Figure S2 Plasma GFAP does not significantly mediate or moderate the associations between tau pathology and neurodegeneration/clinical severity.**

**a,e**, Mediation models testing the association between tau PET and structural MRI measures (adjusted total hippocampal volume (THV) in the (a) Knight ADRC and (e) ADNI cohorts). **b,f**, Mediation models testing the association between tau PET and clinical severity (CDR-SB) in the (b) Knight ADRC and (f) ADNI cohorts. Solid paths denote statistically significant effects and dashed paths denote non-significant effects. Standardized β coefficients are reported. **c,g**, Scatter plots showing the associations between tau PET and structural MRI measures according to plasma GFAP levels in the (c) Knight ADRC and (g) ADNI cohorts. **d,h**, Scatter plots showing the associations between tau PET and clinical severity (CDR-SB) according to plasma GFAP levels in the (d) Knight ADRC and (h) ADNI cohorts. β estimates and corresponding two-sided P values were computed from linear regression models. Shaded areas represent 95% confidence intervals of the regression lines. The P values for were adjusted using Benjamini–Hochberg false discovery rate (BH-FDR) correction, and the corrected P values are denoted as P_FDR_. Both unadjusted P and P_FDR_ values are reported. All analyses were adjusted for age, sex, and cognitive status, except those involving CDR-SB, which were adjusted for age and sex only.

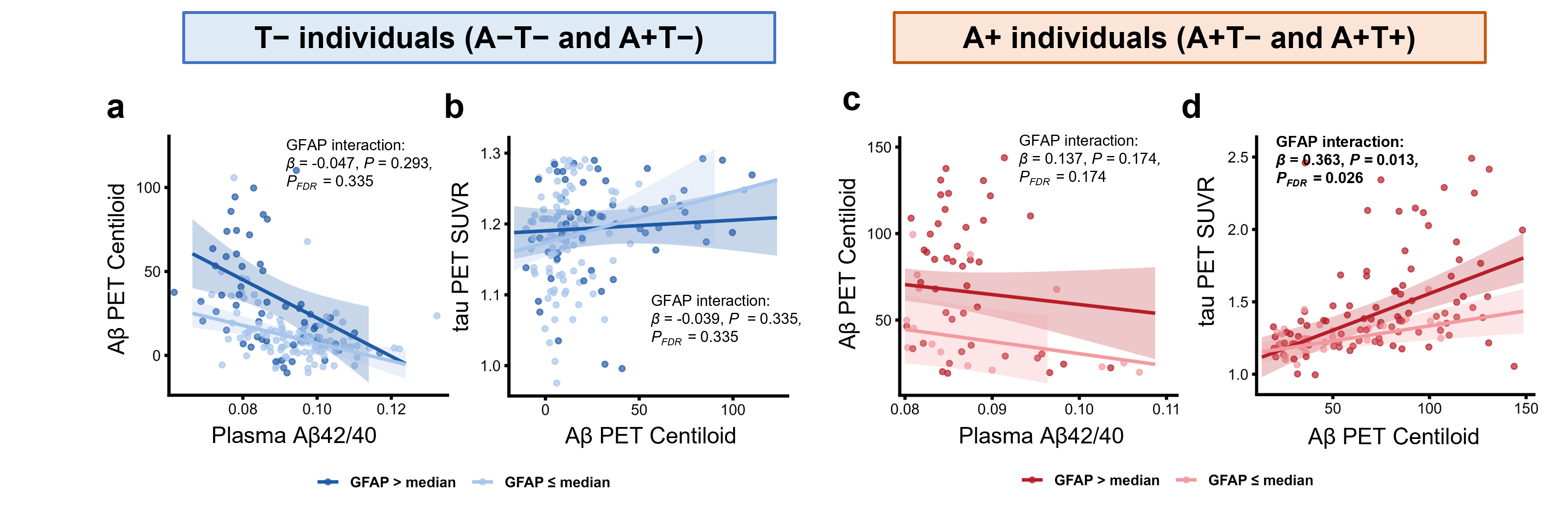

**Figure S3 Moderating roles of astrocytic reactivity with amyloid and tau pathology in A/T-defined subgroups of the ADNI cohort.**

**a,b**, Scatter plots displaying the associations (a) between plasma Aβ42/40 and Aβ PET burden, and (b) between Aβ PET and tau PET according to plasma GFAP levels in T− individuals. **c,d**, Scatter plots displaying the associations (c) between plasma Aβ42/40 and Aβ PET burden, and (d) between Aβ PET and tau PET according to plasma GFAP levels in A+ individuals. β estimates and corresponding two-sided P values were computed from linear regression models. Shaded areas represent 95% confidence intervals of the regression lines. The P values for were adjusted using Benjamini–Hochberg false discovery rate (BH-FDR) correction, and the corrected P values are denoted as P_FDR_. Both unadjusted P and P_FDR_ values are reported. All analyses were adjusted for age, sex, and cognitive status.

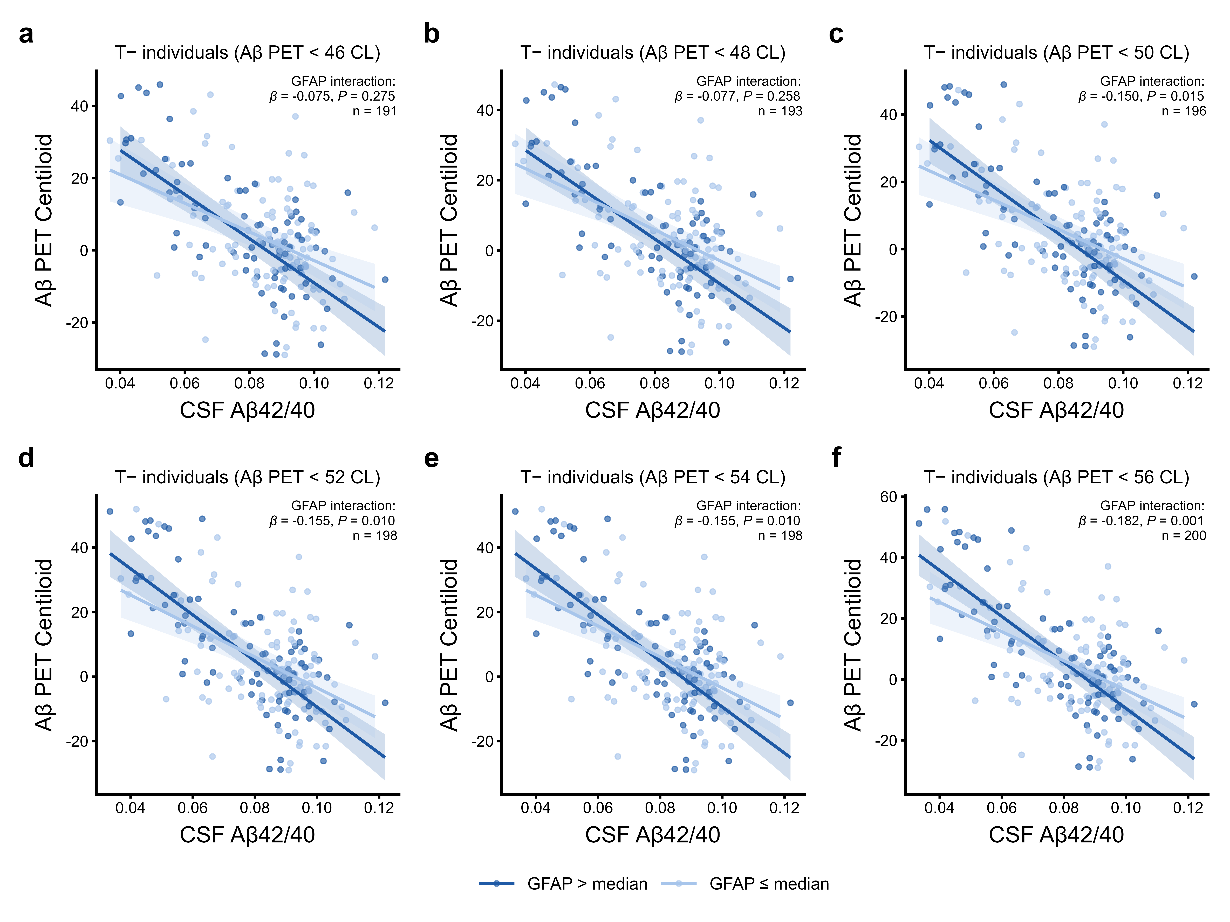
**Figure S4 Sensitivity analyses: plasma GFAP × CSF Aβ42/40 interaction on Aβ PET in tau-negative individuals with relatively lower amyloid burden across Centiloid thresholds.**

a–f, Scatter plots displaying the associations between CSF Aβ42/40 and Aβ PET burden among tau PET-negative (T−) individuals with Aβ PET values below 46 (a), 48 (b), 50 (c), 52 (d), 54 (e), and 56 (f) Centiloids, respectively. The GFAP × CSF Aβ42/40 interaction reached statistical significance at thresholds of 50 Centiloids and above (all P ≤ 0.015). Solid lines represent fitted regression lines, and shaded areas indicate 95% confidence intervals. β estimates and corresponding two-sided P values were computed from linear regression models. Shaded areas represent 95% confidence intervals of the regression lines. All analyses were adjusted for age, sex, and cognitive status.

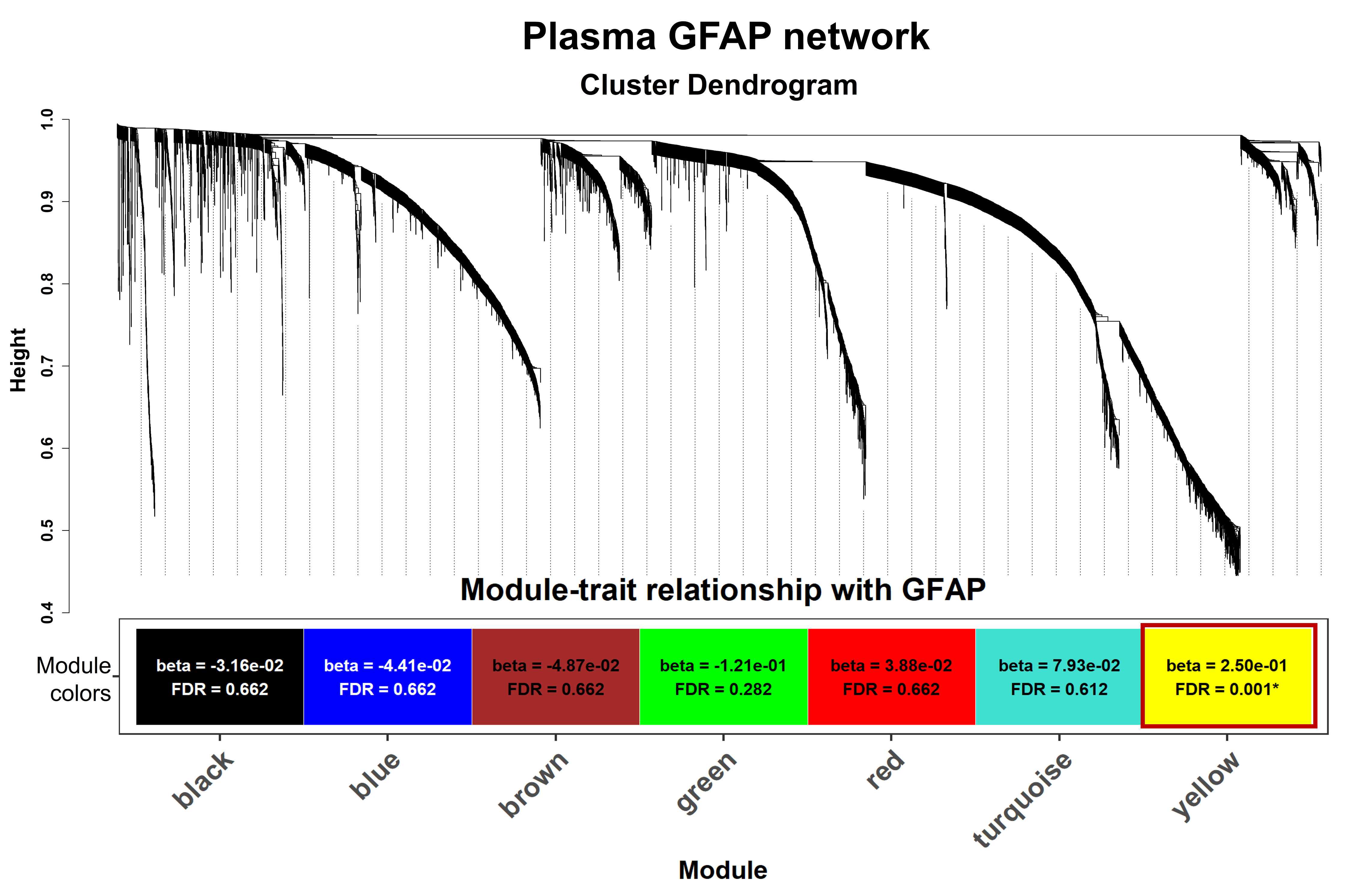

**Figure S5 Sensitivity analysis excluding GFAP from the WGCNA input.**

Cluster dendrogram of plasma proteomic profiles generated by weighted gene co-expression network analysis (WGCNA) repeated after removing GFAP, with proteins grouped into distinct co-expression modules. Heatmap showing correlations between module eigengenes and plasma GFAP. The yellow module (M4) was significantly associated with plasma GFAP after false discovery rate (FDR) correction. β represents the SD change in module eigengene per 1-SD increase in plasma GFAP.

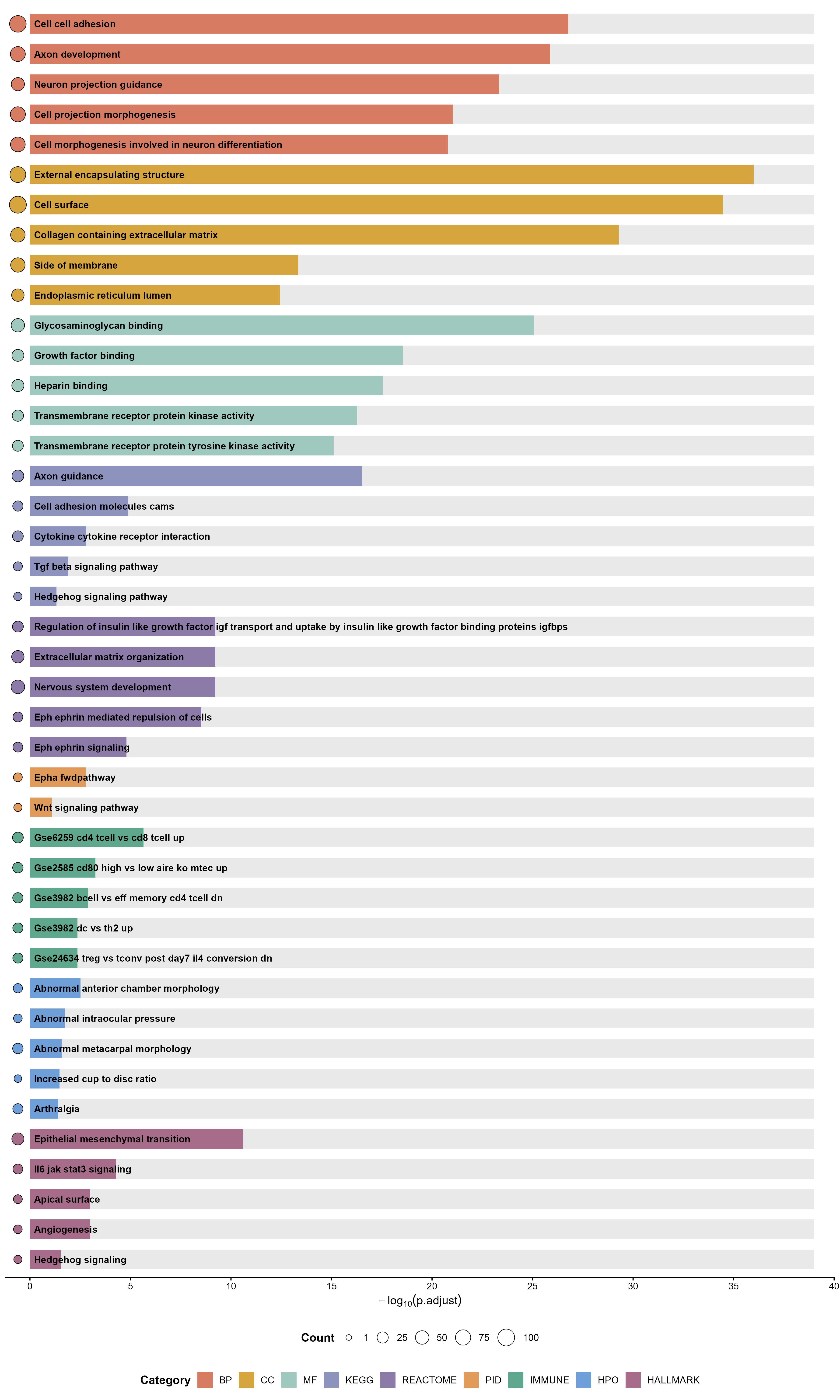

**Figure S6 Functional enrichment analyses of proteins in the MEyellow module.**

Functional enrichment analyses were conducted across GO biological process, cellular component, and molecular function terms, Kyoto Encyclopedia of Genes and Genomes, Reactome, Pathway Interaction Database, Hallmark, ImmuneSigDB, and Human Phenotype Ontology gene sets. The top five significantly enriched terms or pathways with FDR-corrected P < 0.05 from each annotation database are shown. BP, biological process; CC, cellular component; GO, Gene Ontology; HPO, Human Phenotype Ontology; IMMUNE, ImmuneSigDB; KEGG, Kyoto Encyclopedia of Genes and Genomes; MF, molecular function; PID, Pathway Interaction Database.

Table S1 Participant characteristics by A/T status in Knight ADRC cohort.

| Groups | Joint A/T groups | | | Subgroups | |
| --- | --- | --- | --- | --- | --- |
|  | **A-T-** | **A+T-** | **A+T+** | **T-** | **A+** |
| n | 164 | 77 | 37 | 241 | 114 |
| Age, years | 67.5 (8.7) | 71.9 (7.0) | 72.7 (8.9) | 68.9 (8.4) | 72.2 (7.6) |
| Education (years) | 16.5 (2.4) | 16.4 (2.4) | 15.4 (2.6) | 16.5 (2.4) | 16.1 (2.5) |
| Female (no., %) | 84, 51.2% | 44, 57.1% | 16, 43.2% | 128, 53.1% | 60, 52.6% |
| APOE ε4 (no., %) | 49, 29.9% | 47, 61.0% | 22, 59.5% | 96, 39.8% | 69, 60.5% |
| Cognitive diagnosis (no., %) |  |  |  |  |  |
| CN | 164, 100.0% | 63, 81.8% | 2, 5.4% | 227, 94.2% | 65, 57.0% |
| MCI | 0, 0% | 14, 18.2% | 20, 54.1% | 14, 5.8% | 34, 29.8% |
| AD | 0, 0% | 0, 0% | 15, 40.5% | 0, 0% | 15, 13.2% |
| CDR-SB | 0.00 (0.00) | 0.36 (0.87) | 3.89 (2.24) | 0.12 (0.52) | 1.51 (2.20) |
| Plasma GFAP, pg/mL (log_10_-transformed) | 2.08 (0.22) | 2.23 (0.19) | 2.41 (0.15) | 2.13 (0.22) | 2.29 (0.20) |
| Aβ PET, Centiloid (CL) | -0.5 (11.1) | 54.9 (21.6) | 99.3 (41.3) | 17.2 (30.0) | 69.3 (36.0) |
| Tau PET, temporal meta-ROI SUVR | 0.76 (0.13) | 0.89 (0.15) | 2.23 (0.80) | 0.80 (0.14) | 1.33 (0.78) |

Data are presented as mean (SD) for continuous variables and n (%) for categorical variables. The T− subgroup comprises A−T− and A+T− individuals; the A+ subgroup comprises A+T− and A+T+ individuals.

Table S2 Participant characteristics by A/T status in ADNI cohort.

| Groups | Joint A/T groups | | | Subgroups | |
| --- | --- | --- | --- | --- | --- |
|  | **A-T-** | **A+T-** | **A+T+** | **T-** | **A+** |
| n | 113 | 54 | 69 | 167 | 123 |
| Age, years | 69.6 (5.8) | 72.9 (7.8) | 73.8 (7.2) | 70.6 (6.7) | 73.4 (7.5) |
| Education (years) | 16.8 (2.1) | 16.4 (2.4) | 16.0 (2.5) | 16.7 (2.2) | 16.2 (2.4) |
| Female (no., %) | 67, 59.3% | 31, 57.4% | 33, 47.8% | 98, 58.7% | 64, 52.0% |
| APOE ε4 (no., %) | 26, 23.0% | 26, 48.1% | 49, 71.0% | 52, 31.1% | 75, 61.0% |
| Cognitive diagnosis (no., %) |  |  |  |  |  |
| CN | 113, 100.0% | 35, 64.8% | 15, 21.7% | 148, 88.6% | 50, 40.7% |
| MCI | 0, 0.0% | 15, 27.8% | 30, 43.5% | 15, 9.0% | 45, 36.6% |
| AD | 0, 0.0% | 4, 7.4% | 24, 34.8% | 4, 2.4% | 28, 22.8% |
| CDR-SB | 0.06 (0.18) | 0.83 (1.47) | 2.03 (1.92) | 0.31 (0.92) | 1.50 (1.83) |
| Plasma GFAP, pg/mL (log_10_-transformed) | 2.02 (0.21) | 2.19 (0.21) | 2.31 (0.15) | 2.08 (0.22) | 2.26 (0.19) |
| Aβ PET, Centiloid (CL) | 6.0 (7.3) | 48.9 (30.1) | 84.1 (29.9) | 19.9 (27.0) | 68.7 (34.6) |
| Tau PET, temporal meta-ROI SUVR | 1.18 (0.07) | 1.20 (0.07) | 1.61 (0.35) | 1.18 (0.07) | 1.43 (0.34) |

Data are presented as mean (SD) for continuous variables and n (%) for categorical variables. The T− subgroup comprises A−T− and A+T− individuals; the A+ subgroup comprises A+T− and A+T+ individuals.

Table S3 Time differences between imaging, fluid, and clinical assessments in the Knight ADRC and ADNI cohorts.

| **Knight ADRC** | **Median (IQR) time difference, days** | **ADNI** | **Median (IQR) time difference, days** |
| --- | --- | --- | --- |
| CSF Aβ42/40 and Plasma GFAP | 0 (0) | Plasma Aβ42/40 and Plasma GFAP | 0 (0) |
| CSF Aβ42/40 and Aβ PET | 36 (66) | Plasma Aβ42/40 and Aβ PET | 1.5 (15.25) |
| CSF Aβ42/40 and tau PET | 31 (62) | Plasma Aβ42/40 and tau PET | 13 (39) |
| CSF Aβ42/40 and MRI | 34 (64) | Plasma Aβ42/40 and MRI | 33.5 (43) |
| CSF Aβ42/40 and CDR-SB | 73 (67) | Plasma Aβ42/40 and CDR-SB | 4 (18) |
| Plasma GFAP and Aβ PET | 36 (66) | Plasma GFAP and Aβ PET | 1.5 (15.25) |
| Plasma GFAP and tau PET | 31 (62) | Plasma GFAP and tau PET | 13 (39) |
| Plasma GFAP and MRI | 34 (64) | Plasma GFAP and MRI | 33.5 (43) |
| Plasma GFAP and CDR-SB | 73 (67) | Plasma GFAP and CDR-SB | 4 (18) |
| Aβ PET and tau PET | 25 (54) | Aβ PET and tau PET | 9.5 (22.25) |
| Aβ PET and MRI | 0 (0) | Aβ PET and MRI | 31 (34) |
| Aβ PET and CDR-SB | 85 (77) | Aβ PET and CDR-SB | 0 (3) |
| tau PET and MRI | 22 (49) | tau PET and MRI | 42.5 (51.75) |
| tau PET and CDR-SB | 99 (74) | tau PET and CDR-SB | 14 (35) |
| MRI and CDR-SB | 88 (73) | MRI and CDR-SB | 23 (25) |

Table S4 Participant characteristics of the Knight ADRC cohort according to availability of plasma proteomics data.

|  | With plasma proteomics | Without plasma proteomics |
| --- | --- | --- |
| n | 252 | 26 |
| Age, years | 69.6 (8.2) | 68.0 (11.6) |
| Education (years) | 16.3 (2.5) | 16.2 (2.4) |
| Female (no., %) | 128, 50.8% | 16, 61.5% |
| APOE ε4 (no., %) | 108, 42.9% | 10, 38.5% |
| Cognitive diagnosis (no., %) |  |  |
| CN | 209, 82.9% | 20, 76.9% |
| MCI | 28,11.1% | 6, 23.1% |
| AD | 15, 5.9% | 0, 0% |
| CDR-SB | 0.63 (1.64) | 0.52 (1.09) |
| Plasma GFAP, pg/mL (log_10_-transformed) | 2.17 (0.23) | 2.15 (0.27) |
| Aβ PET, Centiloid (CL) | 28.6 (42.6) | 23.6 (38.4) |
| Tau PET, temporal meta-ROI SUVR | 0.99 (0.59) | 1.00 (0.41) |
| Aβ PET (+) (no., %) | 103, 40.9% | 11, 42.3% |
| tau PET (+) (no., %) | 32, 12.7% | 5, 19.2% |

Data are presented as mean (SD) for continuous variables and n (%) for categorical variables.

Table S5 Mediation effects of plasma GFAP for the association between different components of the AD cascade.

| **Cohort** | **Effect** | **Association** | **Coefficients** | **95%CI** | ***P*-value** | ***P*-value**  **(after FDR)** |
| --- | --- | --- | --- | --- | --- | --- |
| **Knight ADRC** | Total | CSF Aβ42/40 → Aβ PET | -0.568 | [-0.644, -0.478] | <0.001 | – |
|  |  | Aβ PET → tau PET | 0.445 | [0.311, 0.601] | <0.001 | – |
|  |  | tau PET → THV | -0.171 | [-0.310, -0.038] | 0.007 | – |
|  |  | tau PET → CDR-SB | 0.716 | [0.594, 0.810] | <0.001 | – |
|  | Direct | CSF Aβ42/40 → Aβ PET | -0.529 | [-0.608, -0.430] | <0.001 | – |
|  |  | Aβ PET → tau PET | 0.397 | [0.260, 0.548] | <0.001 | – |
|  |  | tau PET → THV | -0.157 | [-0.300, -0.021] | 0.017 | – |
|  |  | tau PET → CDR-SB | 0.692 | [0.563, 0.800] | <0.001 | – |
|  | Indirect | CSF Aβ42/40 → GFAP → Aβ-PET | -0.039 | [-0.074, -0.020] | **0.003** | **0.012** |
|  |  | Aβ PET → GFAP → tau PET | 0.048 | [0.016, 0.094] | **0.013** | **0.027** |
|  |  | tau PET → GFAP → THV | -0.013 | [-0.053, 0.022] | 0.489 | 0.489 |
|  |  | tau PET → GFAP → CDR-SB | 0.023 | [-0.005, 0.059] | 0.139 | 0.185 |
| **ADNI** | Total | Plasma Aβ42/40 → Aβ PET | -0.246 | [-0.335, -0.156] | <0.001 | – |
|  |  | Aβ PET → tau PET | 0.479 | [0.258, 0.626] | <0.001 | – |
|  |  | tau PET → THV | -0.300 | [-0.448, -0.144] | <0.001 | – |
|  |  | tau PET → CDR-SB | 0.413 | [0.260, 0.560] | <0.001 | – |
|  | Direct | Plasma Aβ42/40 → Aβ PET | -0.202 | [-0.293, -0.109] | <0.001 | – |
|  |  | Aβ PET → tau PET | 0.417 | [0.202, 0.565] | <0.001 | – |
|  |  | tau PET → THV | -0.277 | [-0.429, -0.120] | <0.001 | – |
|  |  | tau PET → CDR-SB | 0.376 | [0.217, 0.536] | <0.001 | – |
|  | Indirect | Plasma Aβ42/40 → GFAP → Aβ PET | -0.044 | [-0.082, -0.019] | **0.005** | **0.010** |
|  |  | Aβ PET → GFAP → tau PET | 0.062 | [0.029, 0.111] | **0.002** | **0.008** |
|  |  | tau PET → GFAP → THV | -0.023 | [-0.061, 0.004] | 0.146 | 0.146 |
|  |  | tau PET → GFAP → CDR-SB | 0.037 | [-0.000, 0.078] | 0.069 | 0.092 |

Mediation models were adjusted for age, sex and cognitive status, except those involving CDR-SB, which were adjusted for age and sex only.

Table S6 Moderation effects of plasma GFAP for the association between different components of the AD cascade

| **Cohort** | **Statistical model** | **β (95% CI)** | ***P*-values** | ***P*-value**  **(after FDR)** |
| --- | --- | --- | --- | --- |
| **Knight ADRC** | **All sample：** **Aβ PET ~** **CSF Aβ42/40 +** **plasma GFAP +** **CSF Aβ42/40****🞨plasma GFAP + age + sex + cognitive status** | | | |
|  | CSF Aβ42/40🞨plasma GFAP | -0.252 (-0.354, -0.150) | **<0.001** | **<0.001** |
|  | **Plasma GFAP > median：****Aβ PET ~** **CSF Aβ42/40 + age + sex + cognitive status** | | | |
|  | CSF Aβ42/40 | -1.197 (-1.376, -1.019) | <0.001 |  |
|  | **Plasma GFAP** **< median：****Aβ PET ~ CSF Aβ42/40 + age + sex + cognitive status** | | | |
|  | CSF Aβ42/40 | -0.643 (-0.755, -0.532) | <0.001 |  |
|  | **All sample：tau PET ~** **Aβ PET + plasma GFAP + Aβ PET🞨plasma GFAP + age + sex + cognitive status** | | | |
|  | Aβ PET 🞨plasma GFAP | 0.152 (0.091, 0.214) | **<0.001** | **<0.001** |
|  | **Plasma GFAP > median：tau PET ~ Aβ PET + age + sex + cognitive status** | | | |
|  | Aβ PET | 0.733 (0.610, 0.856) | <0.001 |  |
|  | **Plasma GFAP < median：tau PET ~ Aβ PET + age + sex + cognitive status** | | | |
|  | Aβ PET | 0.098 (0.034, 0.162) | 0.003 |  |
|  | **All sample：THV ~ tau PET + plasma GFAP + tau PET🞨plasma GFAP + age + sex + cognitive status** | | | |
|  | tau PET 🞨plasma GFAP | 0.021 (-0.095, 0.137) | 0.722 | 0.722 |
|  | **Plasma GFAP > median：THV ~ tau PET + age + sex + cognitive status** | | | |
|  | tau PET | -0.172 (-0.314, -0.031) | 0.017 |  |
|  | **Plasma GFAP < median：THV ~ tau PET + age + sex + cognitive status** | | | |
|  | tau PET | -0.299 (-0.860, 0.263) | 0.295 |  |
|  | **All sample：****CDR-SB ~** **tau PET + plasma GFAP + tau PET🞨plasma GFAP + age + sex** | | | |
|  | tau PET 🞨plasma GFAP | 0.064 (-0.029, 0.156) | 0.177 | 0.236 |
|  | **Plasma GFAP > median：CDR-SB ~ tau PET + age + sex** | | | |
|  | tau PET | 0.796 (0.690, 0.903) | <0.001 |  |
|  | **Plasma GFAP < median：CDR-SB ~** **tau PET + age + sex** | | | |
|  | tau PET | 0.520 (0.336, 0.704) | <0.001 |  |
| **ADNI** | **All sample： Aβ PET ~ plasma Aβ42/40 +** **plasma GFAP + plasma Aβ42/40🞨plasma GFAP + age + sex + cognitive status** | | | |
|  | plasma Aβ42/40🞨plasma GFAP | -0.031 (-0.122, 0.059) | **0.496** | **0.662** |
|  | **Plasma GFAP > median：Aβ PET ~ plasma Aβ42/40 + age + sex + cognitive status** | | | |
|  | plasma Aβ42/40 | -0.538 (-0.733, -0.343) | <0.001 |  |
|  | **Plasma GFAP < median：Aβ PET ~ plasma Aβ42/40 + age + sex + cognitive status** | | | |
|  | plasma Aβ42/40 | -0.288 (-0.386, -0.190) | <0.001 |  |
|  | **All sample：tau PET ~ Aβ PET + plasma GFAP + Aβ PET🞨plasma GFAP + age + sex + cognitive status** | | | |
|  | Aβ PET 🞨plasma GFAP | 0.277 (0.143, 0.410) | **<0.001** | **<0.001** |
|  | **Plasma GFAP > median：tau PET ~ Aβ PET + age + sex + cognitive status** | | | |
|  | Aβ PET | 0.686 (0.506, 0.867) | <0.001 |  |
|  | **Plasma GFAP < median：tau PET ~ Aβ PET + age + sex + cognitive status** | | | |
|  | Aβ PET | 0.276 (0.191, 0.360) | <0.001 |  |
|  | **All sample：THV ~ tau PET + plasma GFAP + tau PET🞨plasma GFAP + age + sex + cognitive status** | | | |
|  | tau PET 🞨plasma GFAP | 0.086 (-0.086, 0.258) | 0.325 | 0.651 |
|  | **Plasma GFAP > median：THV ~ tau PET + age + sex + cognitive status** | | | |
|  | tau PET | -0.389 (-0.521, -0.257) | <0.001 |  |
|  | **Plasma GFAP < median：THV ~ tau PET + age + sex + cognitive status** | | | |
|  | tau PET | -0.571 (-0.984, -0.158) | 0.007 |  |
|  | **All sample：CDR-SB ~ tau PET + plasma GFAP + tau PET🞨plasma GFAP + age + sex** | | | |
|  | tau PET 🞨plasma GFAP | 0.005 (-0.175, 0.185) | 0.954 | 0.954 |
|  | **Plasma GFAP > median：CDR-SB ~ tau PET + age + sex** | | | |
|  | tau PET | 0.540 (0.388, 0.691) | <0.001 |  |
|  | **Plasma GFAP < median：CDR-SB ~ tau PET + age + sex** | | | |
|  | tau PET | 0.640 (0.342, 0.937) | <0.001 |  |

β-estimates and corresponding statistics were computed from linear regression models. Continuous biomarker levels were standardized prior to model entry.

Table S7 Mediating roles of astrocytic reactivity with amyloid and tau pathology within T− and A+ individuals.

| **Cohort (stage)** | **Effect** | **Association** | **Coefficients** | **95% CI** | ***P*-value** | ***P*-value**  **(after FDR)** |
| --- | --- | --- | --- | --- | --- | --- |
| **Knight ADRC** **(T−)** | | | | | |  |
|  | Total | CSF Aβ42/40 → Aβ PET | -0.732 | [-0.790, -0.657] | <0.001 | – |
|  |  | Aβ PET → tau-PET | 0.321 | [0.200, 0.454] | <0.001 | – |
|  | Direct | CSF Aβ42/40 → Aβ PET | -0.707 | [-0.769, -0.631] | <0.001 | – |
|  |  | Aβ PET → tau-PET | 0.295 | [0.174, 0.430] | <0.001 | – |
|  | Indirect | CSF Aβ42/40 → GFAP → Aβ PET | -0.025 | [-0.051, -0.008] | **0.022** | **0.044** |
|  |  | Aβ PET → GFAP → tau PET | 0.026 | [-0.010, 0.069] | 0.199 | 0.199 |
| **Knight ADRC** **(A+)** | | | | | |  |
|  | Total | CSF Aβ42/40 → Aβ PET | -0.201 | [-0.332, -0.070] | 0.004 | – |
|  |  | Aβ PET → tau-PET | 0.531 | [0.337, 0.686] | <0.001 | – |
|  | Direct | CSF Aβ42/40 → Aβ PET | -0.175 | [-0.307, -0.045] | 0.010 | – |
|  |  | Aβ PET → tau PET | 0.461 | [0.247, 0.640] | <0.001 | – |
|  | Indirect | CSF Aβ42/40 → GFAP → Aβ PET | -0.026 | [-0.077, 0.001] | 0.143 | 0.143 |
|  |  | Aβ PET → GFAP → tau PET | 0.070 | [0.012, 0.156] | **0.045** | 0.090 |
| **ADNI (T−)** | | | | | |  |
|  | Total | Plasma Aβ42/40 → Aβ PET | -0.277 | [-0.400, -0.136] | <0.001 | – |
|  |  | Aβ PET → tau PET | 0.060 | [-0.185, 0.217] | 0.495 | – |
|  | Direct | Plasma Aβ42/40 → Aβ PET | -0.239 | [-0.357, -0.095] | <0.001 | – |
|  |  | Aβ PET → tau PET | 0.075 | [-0.174, 0.235] | 0.409 | – |
|  | Indirect | Plasma Aβ42/40 → GFAP → Aβ PET | -0.037 | [-0.087, -0.009] | **0.047** | 0.094 |
|  |  | Aβ PET → GFAP → tau PET | -0.015 | [-0.072, 0.021] | 0.512 | 0.512 |
| **ADNI (A+)** | | | | | |  |
|  | Total | Plasma Aβ42/40 → Aβ PET | -0.093 | [-0.240, 0.086] | 0.276 | – |
|  |  | Aβ PET → tau-PET | 0.414 | [0.183, 0.564] | <0.001 | – |
|  | Direct | Plasma Aβ42/40 → Aβ PET | -0.055 | [-0.202, 0.118] | 0.512 | – |
|  |  | Aβ PET → tau PET | 0.349 | [0.122, 0.497] | <0.001 | – |
|  | Indirect | Plasma Aβ42/40 → GFAP → Aβ PET | -0.037 | [-0.101, 0.002] | 0.118 | 0.118 |
|  |  | Aβ PET → GFAP → tau PET | 0.065 | [0.021, 0.134] | **0.021** | **0.042** |

Mediation models were adjusted for age, sex and cognitive status. P values are unadjusted. FDR-adjusted P values (Benjamini–Hochberg) were calculated for the indirect (mediation) effects only, across the two indirect-effect tests within each cohort and stage. Total and direct effects are reported for descriptive purposes and were not corrected.

Table S8 Structural equation model coefficients associated statistics for Fig. 2 e and k. Reported are standardized β (95% CI) P-value and FDR-adjusted p-values.

| **Cohort (stage)** | **Std. β (95% CI)** | ***P*-value** | ***P*-value**  **(after FDR)** |
| --- | --- | --- | --- |
| **Knight ADRC (T−)** | | | |
| **CDR-SB** |  |  |  |
| THV | 0.003 (-0.098, 0.104) | 0.959 | 0.969 |
| ... tau PET | 0.032 (-0.074, 0.138) | 0.557 | 0.819 |
| ... Aβ PET | -0.005 (-0.110, 0.099) | 0.922 | 0.969 |
| ... GFAP | -0.013 (-0.086, 0.061) | 0.735 | 0.954 |
| ... CSF Aβ42/40 | -0.012 (-0.107, 0.083) | 0.801 | 0.969 |
| **THV** |  |  |  |
| tau PET | -0.088 (-0.228, 0.052) | 0.220 | 0.391 |
| ... Aβ PET | -0.004 (-0.212, 0.203) | 0.967 | 0.969 |
| ... GFAP | -0.067 (-0.229, 0.095) | 0.419 | 0.671 |
| ... CSF Aβ42/40 | -0.004 (-0.185, 0.178) | 0.969 | 0.969 |
| **tau PET** |  |  |  |
| Aβ PET | 0.280 (0.088, 0.472) | 0.004 | 0.011 |
| ... GFAP | 0.099 (-0.039, 0.238) | 0.159 | 0.317 |
| ... CSF Aβ42/40 | -0.019 (-0.186, 0.148) | 0.823 | 0.969 |
| **Aβ PET** |  |  |  |
| GFAP | 0.113 (0.036, 0.191) | 0.004 | 0.011 |
| ... CSF Aβ42/40 | -0.707 (-0.773, -0.641) | <0.001 | <0.001 |
| **GFAP** |  |  |  |
| CSF Aβ42/40 | -0.218 (-0.323, -0.113) | <0.001 | <0.001 |
| **ADNI (T−)** |  |  |  |
| **CDR-SB** |  |  |  |
| THV | -0.110 (-0.224, 0.003) | 0.057 | 0.141 |
| ... tau PET | -0.075 (-0.172, 0.022) | 0.131 | 0.262 |
| ... Aβ PET | 0.071 (-0.075, 0.216) | 0.340 | 0.494 |
| ... GFAP | -0.024 (-0.130, 0.081) | 0.652 | 0.745 |
| ... plasma Aβ42/40 | -0.075 (-0.175, 0.025) | 0.140 | 0.263 |
| **THV** |  |  |  |
| tau PET | -0.091 (-0.214, 0.033) | 0.149 | 0.265 |
| ... Aβ PET | 0.129 (-0.011, 0.270) | 0.072 | 0.164 |
| ... GFAP | -0.093 (-0.226, 0.041) | 0.173 | 0.291 |
| ... plasma Aβ42/40 | 0.019 (-0.087, 0.125) | 0.728 | 0.804 |
| **tau PET** |  |  |  |
| Aβ PET | 0.070 (-0.143, 0.283) | 0.519 | 0.664 |
| ... GFAP | -0.072 (-0.257, 0.113) | 0.447 | 0.600 |
| ... plasma Aβ42/40 | -0.018 (-0.199, 0.164) | 0.848 | 0.905 |
| **Aβ PET** |  |  |  |
| GFAP | 0.145 (0.039, 0.250) | 0.007 | 0.021 |
| ... plasma Aβ42/40 | -0.239 (-0.361, -0.117) | <0.001 | <0.001 |
| **GFAP** |  |  |  |
| plasma Aβ42/40 | -0.258 (-0.396, -0.120) | <0.001 | <0.001 |

P values adjusted for multiple comparisons using false discovery rate were considered significant at p < 0.05. Associations were adjusted for age, sex, and (if not involving CDR-SB) cognitive status. In Knight ADRC, the model fit is mixed overall (n = 241, X^2^ = 3.573, degrees of freedom = 1, RMSEA = 0.097, SRMR = 0.007, CFI = 0.997). In ADNI, the model fit the data well (n = 167, X^2^ = 0.119, degrees of freedom = 1, RMSEA = 0.000, SRMR = 0.002, CFI = 1.000).

Table S9 Structural equation model coefficients associated statistics for Fig. 2 f and l. Reported are standardized β (95% CI) P-value and FDR-adjusted p-values.

| **Cohort (stage)** | **Std. β (95% CI)** | ***P*-value** | ***P*-value**  **(after FDR)** |
| --- | --- | --- | --- |
| **Knight ADRC (A+)** | | | |
| **CDR-SB** |  |  |  |
| THV | -0.309 (-0.453, -0.164) | <0.001 | <0.001 |
| ... tau PET | 0.459 (0.218, 0.700) | <0.001 | <0.001 |
| ... Aβ PET | 0.122 (-0.121, 0.365) | 0.324 | 0.398 |
| ... GFAP | 0.059 (-0.067, 0.185) | 0.360 | 0.427 |
| ... CSF Aβ42/40 | 0.010 (-0.096, 0.115) | 0.860 | 0.860 |
| **THV** |  |  |  |
| tau PET | -0.451 (-0.687, -0.216) | <0.001 | <0.001 |
| ... Aβ PET | 0.073 (-0.170, 0.315) | 0.557 | 0.614 |
| ... GFAP | -0.093 (-0.266, 0.080) | 0.292 | 0.389 |
| ... CSF Aβ42/40 | -0.090 (-0.217, 0.037) | 0.164 | 0.250 |
| **tau PET** |  |  |  |
| GFAP | 0.162 (0.025, 0.299) | 0.020 | 0.043 |
| ... Aβ PET | 0.466 (0.272, 0.661) | <0.001 | <0.001 |
| ... CSF Aβ42/40 | 0.016 (-0.083, 0.116) | 0.746 | 0.796 |
| **GFAP** |  |  |  |
| Aβ PET | 0.409 (0.236, 0.583) | <0.001 | <0.001 |
| CSF Aβ42/40 | -0.078 (-0.214, 0.059) | 0.265 | 0.369 |
| **Aβ PET** |  |  |  |
| CSF Aβ42/40 | -0.285 (-0.444, -0.127) | <0.001 | 0.001 |
| **ADNI (A+)** |  |  |  |
| **CDR-SB** |  |  |  |
| THV | -0.439 (-0.593, -0.286) | <0.001 | <0.001 |
| ... tau PET | 0.167 (-0.027, 0.361) | 0.091 | 0.154 |
| GFAP | 0.164 (-0.028, 0.355) | 0.094 | 0.154 |
| ... Aβ PET | 0.020 (-0.158, 0.199) | 0.824 | 0.879 |
| ... plasma Aβ42/40 | -0.096 (-0.227, 0.035) | 0.150 | 0.228 |
| **THV** |  |  |  |
| tau PET | -0.308 (-0.511, -0.106) | 0.003 | 0.008 |
| GFAP | -0.050 (-0.235, 0.135) | 0.597 | 0.658 |
| ... Aβ PET | -0.080 (-0.199, 0.038) | 0.185 | 0.261 |
| ... plasma Aβ42/40 | -0.007 (-0.117, 0.102) | 0.896 | 0.896 |
| **tau PET** |  |  |  |
| GFAP | 0.284 (0.170, 0.399) | <0.001 | <0.001 |
| ... Aβ PET | 0.358 (0.175, 0.541) | <0.001 | <0.001 |
| ... plasma Aβ42/40 | 0.141 (0.033, 0.249) | 0.010 | 0.026 |
| **GFAP** |  |  |  |
| Aβ PET | 0.234 (0.079, 0.389) | 0.003 | 0.008 |
| ... plasma Aβ42/40 | -0.150 (-0.311, 0.012) | 0.069 | 0.130 |
| **Aβ PET** |  |  |  |
| plasma Aβ42/40 | -0.093 (-0.251, 0.066) | 0.251 | 0.335 |

P values adjusted for multiple comparisons using false discovery rate were considered significant at p < 0.05. Associations were adjusted for age, sex, and (if not involving CDR-SB) cognitive status. In Knight ADRC, the model fit the data well (n = 114, X^2^ = 1.066, degrees of freedom = 1, RMSEA = 0.009, SRMR = 0.006, CFI = 1.000). In ADNI, the model fit the data well (n = 123, X^2^ = 1.029, degrees of freedom = 1, RMSEA = 0.006, SRMR = 0.007, CFI = 1.000).

Table S10 Moderating roles of astrocytic reactivity with amyloid and tau pathology within T− and A+ individuals.

| **Cohort (stage)** | **Statistical model** | **β (95% CI)** | ***P*-values** | ***P*-value**  **(after FDR)** |
| --- | --- | --- | --- | --- |
| **Knight ADRC (T−)** | **All sample： Aβ PET ~ CSF Aβ42/40 + plasma GFAP + CSF Aβ42/40🞨plasma GFAP + age + sex + cognitive status** | | |  |
|  | CSF Aβ42/40🞨plasma GFAP | -0.180 (-0.261, -0.100) | **<0.001** | **<0.001** |
|  | **Plasma GFAP > median：Aβ PET ~ CSF Aβ42/40 + age + sex + cognitive status** | | |  |
|  | CSF Aβ42/40 | -0.873 (-0.999, -0.748) | <0.001 |  |
|  | **Plasma GFAP < median：Aβ PET ~ CSF Aβ42/40 + age + sex + cognitive status** | | |  |
|  | CSF Aβ42/40 | -0.523 (-0.636, -0.410) | <0.001 |  |
|  | **All sample：tau PET ~ Aβ PET + plasma GFAP + Aβ PET🞨plasma GFAP + age + sex + cognitive status** | | |  |
|  | Aβ PET 🞨plasma GFAP | 0.037 (-0.083, 0.157) | 0.544 | 0.544 |
|  | **Plasma GFAP > median：tau PET ~ Aβ PET + age + sex + cognitive status** | | |  |
|  | Aβ PET | 0.358 (0.210, 0.507) | <0.001 |  |
|  | **Plasma GFAP < median：tau PET ~ Aβ PET + age + sex + cognitive status** | | |  |
|  | Aβ PET | 0.180 (-0.030, 0.389) | 0.092 |  |
| **Knight ADRC (A+)** | **All sample：Aβ PET ~ CSF Aβ42/40 + plasma GFAP + CSF Aβ42/40🞨plasma GFAP + age + sex + cognitive status** | | |  |
|  | CSF Aβ42/40🞨plasma GFAP | -0.059 (-0.211, 0.093) | 0.44 | 0.44 |
|  | **Plasma GFAP > median：Aβ PET ~ CSF Aβ42/40 + age + sex + cognitive status** | | |  |
|  | CSF Aβ42/40 | -0.322 (-0.584, -0.060) | 0.017 |  |
|  | **Plasma GFAP < median：Aβ PET ~ CSF Aβ42/40 + age + sex + cognitive status** | | |  |
|  | CSF Aβ42/40 | -0.108 (-0.234, 0.018) | 0.089 |  |
|  | **All sample：tau PET ~ Aβ PET + plasma GFAP + Aβ PET🞨plasma GFAP + age + sex + cognitive status** | | |  |
|  | Aβ PET 🞨plasma GFAP | 0.108 (-0.005, 0.221) | **0.061** | **0.122** |
|  | **Plasma GFAP > median：tau PET ~ Aβ PET + age + sex + cognitive status** | | |  |
|  | Aβ PET | 0.488 (0.303, 0.673) | <0.001 |  |
|  | **Plasma GFAP < median：tau PET ~ Aβ PET + age + sex + cognitive status** | | |  |
|  | Aβ PET | 0.136 (-0.046, 0.319) | 0.135 |  |
| **ADNI (T−)** | **All sample： Aβ PET ~ plasma Aβ42/40 + plasma GFAP + plasma Aβ42/40🞨plasma GFAP + age + sex + cognitive status** | | |  |
|  | plasma Aβ42/40🞨plasma GFAP | -0.047 (-0.135, 0.041) | 0.293 | 0.335 |
|  | **Plasma GFAP > median：Aβ PET ~ plasma Aβ42/40 + age + sex + cognitive status** | | |  |
|  | plasma Aβ42/40 | −0.337 (-0.567, -0.108) | 0.005 |  |
|  | **Plasma GFAP < median：Aβ PET ~ plasma Aβ42/40 + age + sex + cognitive status** | | |  |
|  | plasma Aβ42/40 | −0.156 (-0.235, -0.077) | < 0.001 |  |
|  | **All sample：tau PET ~ Aβ PET + plasma GFAP + Aβ PET🞨plasma GFAP + age + sex + cognitive status** | | |  |
|  | Aβ PET 🞨plasma GFAP | -0.039 (-0.119, 0.041) | 0.335 | 0.335 |
|  | **Plasma GFAP > median：tau PET ~ Aβ PET + age + sex + cognitive status** | | |  |
|  | Aβ PET | 0.022 (-0.057, 0.101) | 0.583 |  |
|  | **Plasma GFAP < median：tau PET ~ Aβ PET + age + sex + cognitive status** | | |  |
|  | Aβ PET | 0.105 (-0.013, 0.222) | 0.081 |  |
| **ADNI (A+)** | **All sample：Aβ PET ~ plasma Aβ42/40 + plasma GFAP + CSF Aβ42/40🞨plasma GFAP + age + sex + cognitive status** | | |  |
|  | plasma Aβ42/40 🞨plasma GFAP | 0.137 (-0.061, 0.335) | 0.174 | 0.174 |
|  | **Plasma GFAP > median：Aβ PET ~ plasma Aβ42/40 + age + sex + cognitive status** | | |  |
|  | plasma Aβ42/40 | -0.170 (−0.426, 0.085) | 0.189 |  |
|  | **Plasma GFAP < median：Aβ PET ~ plasma Aβ42/40 + age + sex + cognitive status** | | |  |
|  | plasma Aβ42/40 | −0.208 (−0.450, 0.035) | 0.090 |  |
|  | **All sample：tau PET ~ Aβ PET + plasma GFAP + Aβ PET🞨plasma GFAP + age + sex + cognitive status status** | | |  |
|  | Aβ PET 🞨plasma GFAP | 0.363 (0.079, 0.647) | **0.013** | **0.026** |
|  | **Plasma GFAP > median：tau PET ~ Aβ PET + age + sex + cognitive status** | | |  |
|  | Aβ PET | 0.735 (0.460, 1.011) | < 0.001 |  |
|  | **Plasma GFAP < median：tau PET ~ Aβ PET + age + sex + cognitive status** | | |  |
|  | Aβ PET | 0.302 (0.117, 0.487) | 0.002 |  |

β-estimates and corresponding statistics were computed from linear regression models. Continuous biomarker levels were standardized prior to model entry.

Table S11 Associations between plasma GFAP and protein modules.

| **Module** | **Trait** | **Standardized β** | **SE** | **t value** | **P_FDR_** | **Covariates** |
| --- | --- | --- | --- | --- | --- | --- |
| **MEblack** | GFAP | −0.032 | 0.072 | −0.438 | 0.662 | age + sex + PlateId |
| **MEred** | GFAP | 0.039 | 0.068 | 0.567 | 0.662 | age + sex + PlateId |
| **MEturquoise** | GFAP | 0.079 | 0.071 | 1.123 | 0.613 | age + sex + PlateId |
| **MEbrown** | GFAP | −0.049 | 0.072 | −0.681 | 0.662 | age + sex + PlateId |
| **MEgreen** | GFAP | −0.121 | 0.069 | −1.754 | 0.283 | age + sex + PlateId |
| **MEblue** | GFAP | −0.044 | 0.068 | −0.648 | 0.662 | age + sex + PlateId |
| **MEyellow** | GFAP | 0.250 | 0.065 | 3.875 | < 0.001 | age + sex + PlateId |

β coefficients and P values are derived from multivariable linear regression models, controlling for age, sex, and assay plate (PlateId).
